# Indirect effects of doxyPEP implementation reveal core-group dynamics of bacterial sexually-transmitted infections

**DOI:** 10.64898/2026.07.21.26358607

**Authors:** Matan Yechezkel, Banshri Kapadia, Vennis Hong, Jue Tao Lim, Iris Anne C. Reyes, Magdalena E. Pomichowski, Gregg S. Davis, Isabel Rodriguez-Barraquer, Nicola F. Müller, Sara Y. Tartof, Joseph A. Lewnard

## Abstract

Epidemiological theory holds that core groups of high-risk individuals sustain pathogen transmission. However, the scale at which these dynamics operate is poorly understood. We leveraged doxycycline post-exposure prophylaxis (doxyPEP) implementation among at-risk men who have sex with men (MSM) and transgender women (TGW) as a natural experiment to probe bacterial sexually transmitted infection (STI) dynamics within and across sexual networks. DoxyPEP uptake by 9% of at-risk MSM and TGW was associated with pronounced indirect protection within their sexual networks, substantiating the notion that core groups of individuals wield outsize influence on transmission. Risk of chlamydia and syphilis declined by 80% (95% confidence interval: 71–86%) and 65% (30–82%), respectively, among doxyPEP recipients, and by 41% (32–48%) and 29% (13–42%), respectively, among MSM and TGW who did not receive doxyPEP. However, this substantial reduction in disease among at-risk MSM and TGW did not impact incidence among females and other males. Our findings substantiate core group dynamics of STIs among MSM and TGW but call into question the “bisexual bridge” hypothesis positioning this population as a core group impacting STI dynamics among heterosexuals. These findings should inform rational expectations for the impacts of doxyPEP in settings implementing this intervention.

## INTRODUCTION

The “core group” theory in infectious disease posits that small population subsets, distinguished by elevated infection risk, wield outsize influence over transmission of certain pathogens.^1^ Under this framework, contributions of high-risk individuals to transmission permit the basic reproduction number (*R*_0_) to exceed 1 for infections otherwise exhibiting sub-critical transmission among most of the population; simultaneously, sporadic involvement of lower-risk individuals ensures that saturation of infection among high-risk individuals does not curtail transmission. The notion of core groups emerged in studies of the epidemiology of bacterial sexually-transmitted infections (bSTIs),^2–4^ and thereafter provided a valuable framework for understanding dynamics of HIV transmission.^5,6^ Population-level reductions in HIV incidence associated with the targeted deployment of preventive interventions among high-risk individuals substantiate the role of core groups in transmission.^7–9^

Men who have sex with men (MSM) and transgender women (TGW) experience high risk of bSTIs, and have been suggested to act as a core group shaping bSTI transmission within broader heterosexual networks through spillover via men who have sex with men and women (MSMW)—a phenomenon termed the “bisexual bridge” hypothesis.^10–18^ In comparison to a population-wide incidence rate of ∼0.7 bSTI cases per 100 persons in the United States,^19^ rates of 32-75 bSTI episodes per 100 persons have been reported in MSM cohorts selected on behavioral risk, with infections concentrated among individuals who receive recurrent diagnoses.^20–22^ Behaviors including unprotected intercourse with one-time or recurring partners, attendance at sex-on-premises venues or parties, and engagement in sex under the influence of psychoactive drugs (“chemsex”) may define core groups within the broader MSM and TGW populations experiencing the greatest bSTI risk.^23,24^

Post-exposure prophylaxis with 200mg doxycycline (doxyPEP) recently emerged as a strategy to prevent bSTIs, and is now recommended by the World Health Organization (WHO) for MSM and TGW at high risk of bSTI exposure.^25^ Pre-implementation randomized trials and subsequent observational studies have demonstrated that doxyPEP reduces incidence of chlamydia and syphilis by 61-80% among recipients.^26–29^ Whether doxyPEP implementation has also conferred indirect protection among non-recipients remains uncertain. DoxyPEP implementation provides a natural experiment for probing core-group dynamics in bSTIs, as current guidelines are restricted to MSM and TGW, and within these populations target individuals at high bSTI risk based on risk factors such as those listed above.^30^ We assessed impacts of doxyPEP on bSTI incidence in southern California, one of the first settings to implement this intervention. Our analyses leveraged highly-resolved individual-level data on antibiotic prescribing and bSTI testing to distinguish protection arising through direct and indirect pathways.

## RESULTS

### Probing the role of core groups via indirect effects of doxyPEP

Prior to US-wide recommendations for doxyPEP among MSM and TGW, the California Department of Public Health (CDPH) issued clinical guidance for doxyPEP implementation in April, 2023,^31^ following prior local recommendations for doxyPEP issued by the San Francisco Department of Public Health in October, 2022.^32^ Our analyses quantified changes in bSTI incidence within three populations with differing proximity to doxyPEP recipients within their sexual networks. The first—which we defined as “at-risk” MSM and TGW—aimed to proxy individuals with direct exposure to bSTI circulation within majority-male sexual networks. Indirect effects of doxyPEP implementation among non-recipients within this population could support the hypothesis that high-risk individuals receiving doxyPEP represent a core group shaping bSTI transmission among other MSM and TGW (**Figure 1**). The second and third populations were females and males not meeting the “at-risk” designation, respectively, among whom we expected most bSTI exposure to arise within heterosexual networks. Because doxyPEP eligibility extends only to MSM and TGW, evidence of doxyPEP impact among these populations could support the hypothesis that MSM and TGW function as a core group influencing broader population dynamics of bSTIs.

**Figure 1:**
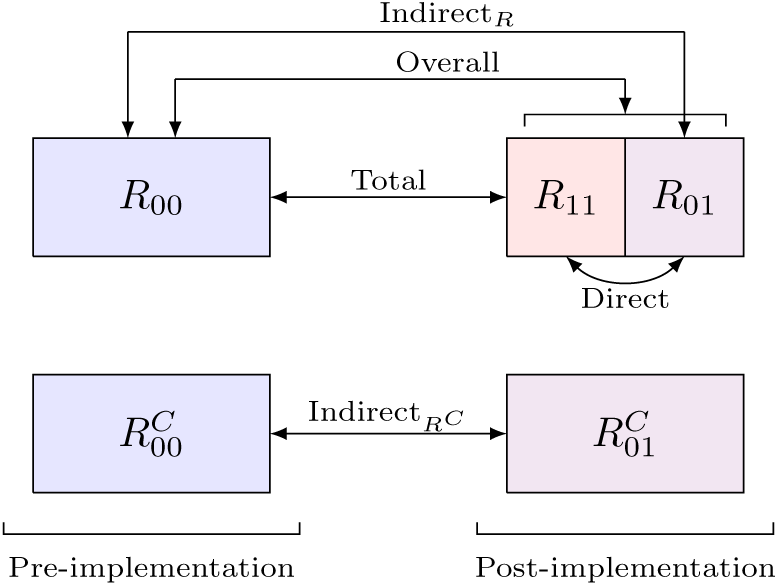
Mapping of counterfactual comparisons and causal estimands. We divide the population across two time periods, before (pre-) and after (post-) implementation of doxyPEP in southern California (January 1, 2022 to April 27, 2023 and April 28, 2023 to June 30, 2025, respectively), and two risk strata, distinguishing at-risk MSM and TGW, *R*, from the remaining (not at-risk) population, *R^C^*. Following the framework of Halloran and colleagues ^36,37^, we define direct effects by comparing outcomes within risk periods 90 days after a doxyPEP dispense for recipients to risk periods not preceded by a doxyPEP dispense within 90 days, within a population exposed to doxyPEP (*R*_11_ and *R*_01_ for exposed and unexposed risk periods within a population with doxyPEP use). We define indirect effects among at-risk MSM and TGW by comparing risk periods not preceded by doxyPEP dispenses, in a doxyPEP-exposed population (*R*_01_) to risk periods not preceded by doxyPEP dispenses in a doxyPEP-unexposed population (*R*_00_). We define indirect effects among the remaining population (*R^C^*) of females and other males by an equivalent comparison across periods with and without population doxyPEP exposure (R^c^_01_ vs. R^c^_00_). We define overall effects among at-risk MSM and TGW by comparing outcomes within risk periods 90 days after a doxyPEP dispense for recipients in a doxyPEP-exposed population (*R*_11_) to risk periods not preceded by doxyPEP dispenses in a doxyPEP-unexposed population (*R*_00_). Last, we define overall effects among at-risk MSM by comparing all risk periods within a doxyPEP-exposed population (*R*_•1_) to risk periods not preceded by doxyPEP dispenses in a doxyPEP-unexposed population (*R*_00_).

To address these questions, we conducted a retrospective cohort study using comprehensive electronic health record (EHR) data from adults aged 16-59 years who belonged to Kaiser Permanente Southern California (KPSC) between July 1, 2021 and June 30, 2025. The KPSC healthcare system provides integrated healthcare services across virtual, outpatient, and hospital settings to ∼5 million individuals throughout southern California, who enroll through employer-sponsored, government-subsidized, and prepaid insurance schemes. Patient EHRs record all diagnoses, prescriptions, procedures, and laboratory tests and results from care delivered at KPSC facilities; further capture of out-of-network care through insurance claim reimbursements enables near-complete ascertainment of members’ healthcare interactions.

### Dynamics of bSTIs among at-risk MSM and TGW

The at-risk MSM and TGW population comprised 30,185 individuals assigned male sex at birth who were living with HIV or receiving HIV pre-or post-exposure prophylaxis (PrEP/PEP) during the study period (**Table S1**; **Figure S1**). Although administrative healthcare data do not enable ascertainment of sexual orientation, MSM and TGW comprise the overwhelming majority of HIV-positive males and nearly all of those receiving PrEP/PEP,^33,34^ and HIV or PrEP/PEP exposures have similarly defined the focal population for past doxyPEP studies.^28,35^ Among this population, 2,713 (9.0%) individuals received ≥1 doxyPEP fill, and 5,967 doxyPEP dispenses, in total, occurred throughout follow-up (**Table S2**). Recipients and non-recipients of doxyPEP resembled each other with respect to race/ethnicity, likelihood of commercial insurance coverage, history of drug/alcohol abuse, and neighborhood socioeconomic characteristics. However, doxyPEP recipients tended to be younger than individuals who never received doxyPEP (median age 33y vs. 35y), less likely to be living with HIV (5.4% vs. 11.1%), more likely to have received JYNNEOS vaccine (65.8% vs. 47.1%), and more likely to have been diagnosed with chlamydia (6.1% vs. 4.1%), gonorrhea (7.0% vs. 4.8%), or syphilis (2.7% vs. 1.8%) within 6 months before study entry.

Following doxyPEP implementation, incidence rates of chlamydia declined from 107.3 to 80.2 cases per 1,000 person-years among all at-risk MSM and TGW, representing a 41.8% (95% confidence interval: 33.2 to 49.4%) adjusted decline against pre-implementation trends in incidence (**Table 1**; **Table 2**; **Figure 2**). For syphilis, incidence rates before and after doxyPEP implementation were 36.1 and 21.3 cases per 1,000 person-years, respectively (29.2% [13.3 to 42.7%] adjusted decline). Corresponding gonorrhea incidence rates before and after doxyPEP implementation were 110.8 and 121.8 cases per 1,000 person-years, respectively (–0.9% [–14.7 to 11.3%] adjusted decline), in agreement with prior evidence that rapid expansions in high-level tetracycline resistance in *N. gonorrhoeae* after doxyPEP implementation precluded meaningful protection against infection (**Table S3**).^27^ Changepoint analyses exploring the timing of changes in incidence trends for each bSTI confirmed that declines in chlamydia and syphilis diagnoses began shortly after doxyPEP implementation, without accompanying changes in gonorrhea incidence rates (**Figure S2**).

**Figure 2:**
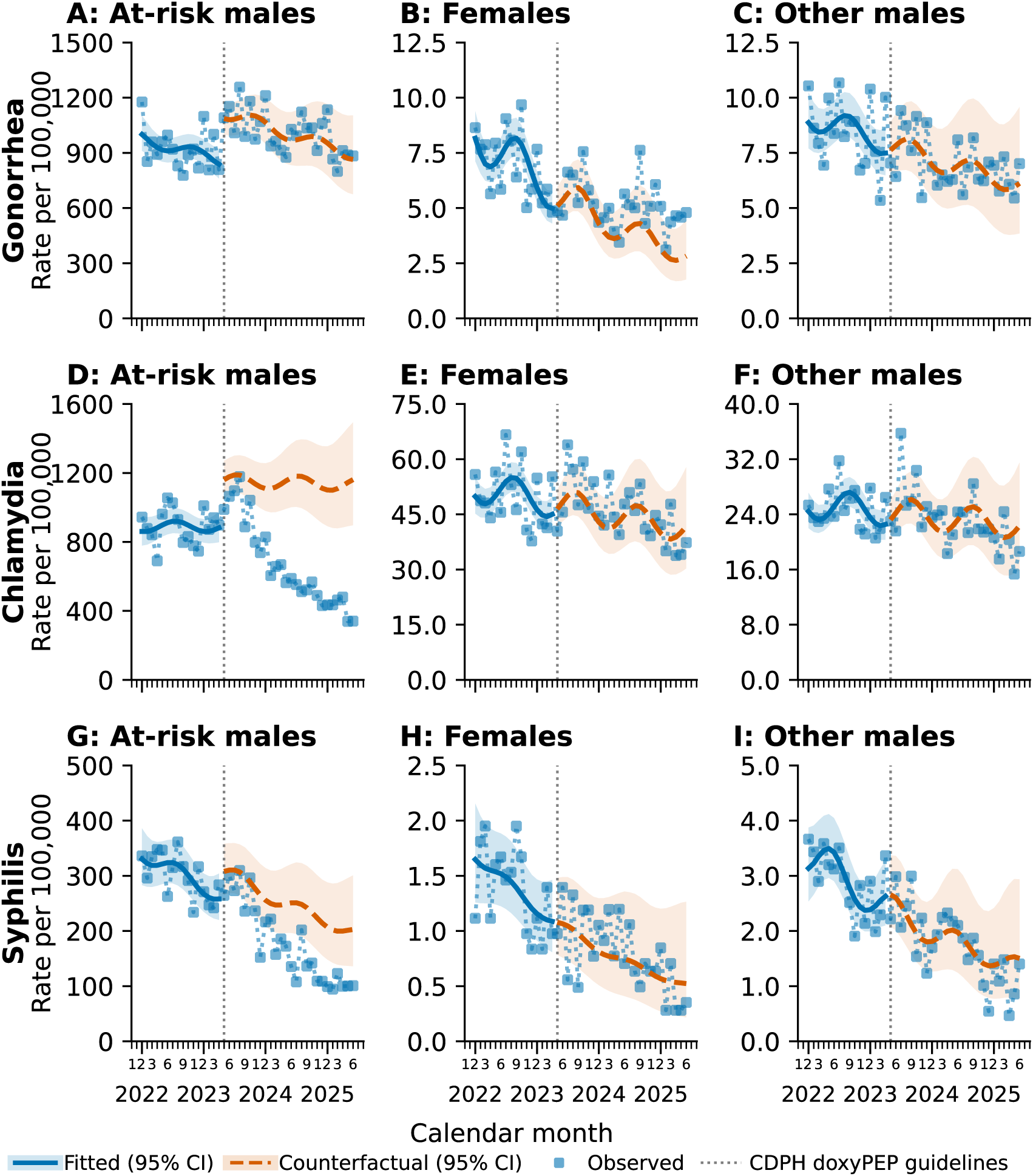
Observed and counterfactual trends in gonorrhea, chlamydia, and syphilis incidence following doxyPEP guideline implementation. We illustrate observed, fitted, and counterfactual monthly incidence rates per 1,000 persons for (a-c) gonorrhea, (d-f) chlamydia, and (g-i) syphilis among (a, d, g) at-risk males (MSM/TGW living with HIV or with a history of receiving HIV PrEP/PEP), (b, e, h) females, and (c, f, i) other males. Counterfactual estimates were obtained using interrupted time-series models with time modeled as a continuous variable and adjustment for age group and seasonal variation. Dashed lines represent the counterfactual estimates, solid lines the fitted model, and points connected by dotted lines the observed data. Shaded areas delineate 95% confidence intervals.

**Table 1:** Incidence rates of bSTI diagnoses and doxycycline dispenses before and after doxyPEP guideline implementation.

| Population | Age group | Period | Incidence rate per 1,000 person-years (95% CI) |  |  |  |
|---|---|---|---|---|---|---|
|  |  |  | Chlamydia, cases<sup>1</sup> | Syphilis, cases<sup>1</sup> | Gonorrhea, cases<sup>1</sup> | Doxycycline, DDDs<sup>2</sup> |
| At-risk MSM and TGW<sup>3</sup> |  |  |  |  |  |  |
|  | All |  |  |  |  |  |
|  |  | January 1, 2022 to April 27, 2023 | 107.3 (103.5 to 111.3) | 36.1 (33.9 to 38.4) | 110.8 (106.9 to 114.8) | 8,204 (7,825 to 8,612) |
|  |  | April 28, 2023 to June 30, 2025 | 80.2 (77.7 to 82.9) | 21.3 (20.0 to 22.7) | 121.8 (118.6 to 125.0) | 27,568 (22,124–32,994) |
|  | 16 to 24y |  |  |  |  |  |
|  |  | January 1, 2022 to April 27, 2023 | 108.1 (97.8 to 119.1) | 29.7 (24.4 to 35.7) | 125.6 (114.5 to 137.4) | 7,498 (6,524 to 8,558) |
|  |  | April 28, 2023 to June 30, 2025 | 111.4 (102.9 to 120.5) | 20.5 (17.0 to 24.6) | 162.3 (152.0 to 173.1) | 12,383 (11,472 to 13,365) |
|  | 25 to 34y |  |  |  |  |  |
|  |  | January 1, 2022 to April 27, 2023 | 142.6 (134.8 to 150.8) | 40.3 (36.1 to 44.7) | 147.2 (139.2 to 155.5) | 9,317 (8,715 to 9,974) |
|  |  | April 28, 2023 to June 30, 2025 | 99.5 (94.5 to 104.6) | 22.5 (20.2 to 25.0) | 160.7 (154.4 to 167.2) | 29,376 (28,406 to 30,375) |
|  | 35 to 44y |  |  |  |  |  |
|  |  | January 1, 2022 to April 27, 2023 | 114.2 (106.4 to 122.4) | 38.5 (34.0 to 43.4) | 120.5 (112.5 to 128.9) | 9,093 (8,309 to 9,949) |
|  |  | April 28, 2023 to June 30, 2025 | 78.8 (73.8 to 84.0) | 23.5 (20.9 to 26.4) | 125.1 (118.8 to 131.6) | 37,226 (35,877 to 38,637) |
|  | 45 to 59y |  |  |  |  |  |
|  |  | January 1, 2022 to April 27, 2023 | 64.4 (59.1 to 70.1) | 32.6 (28.8 to 36.7) | 57.8 (52.8 to 63.2) | 6,615 (5,886 to 7,426) |
|  |  | April 28, 2023 to June 30, 2025 | 46.7 (43.1 to 50.5) | 18.2 (16.0 to 20.7) | 57.7 (53.7 to 61.9) | 23,504 (22,341 to 24,670) |
| Other males |  |  |  |  |  |  |
|  | All |  |  |  |  |  |
|  |  | January 1, 2022 to April 27, 2023 | 3.0 (2.9 to 3.1) | 0.4 (0.3 to 0.4) | 1.0 (1.0 to 1.1) | 1,735 (1,709 to 1,760) |
|  |  | April 28, 2023 to June 30, 2025 | 2.8 (2.7 to 2.8) | 0.2 (0.2 to 0.2) | 0.8 (0.8 to 0.9) | 1,902 (1,842–1,969) |
|  | 16 to 24y |  |  |  |  |  |
|  |  | January 1, 2022 to April 27, 2023 | 7.2 (6.9 to 7.5) | 0.4 (0.3 to 0.4) | 2.2 (2.1 to 2.4) | 3,188 (3,114 to 3,263) |
|  |  | April 28, 2023 to June 30, 2025 | 6.6 (6.4 to 6.8) | 0.2 (0.2 to 0.2) | 1.7 (1.6 to 1.8) | 3,214 (3,150 to 3,279) |
|  | 25 to 34y |  |  |  |  |  |
|  |  | January 1, 2022 to April 27, 2023 | 4.1 (4.0 to 4.3) | 0.5 (0.4 to 0.6) | 1.3 (1.2 to 1.4) | 1,444 (1,402 to 1,488) |
|  |  | April 28, 2023 to June 30, 2025 | 3.8 (3.6 to 3.9) | 0.4 (0.3 to 0.4) | 1.2 (1.1 to 1.2) | 1,573 (1,534 to 1,612) |
|  | 35 to 44y |  |  |  |  |  |
|  |  | January 1, 2022 to April 27, 2023 | 1.5 (1.4 to 1.7) | 0.3 (0.3 to 0.4) | 0.7 (0.6 to 0.8) | 1,303 (1,261 to 1,346) |
|  |  | April 28, 2023 to June 30, 2025 | 1.6 (1.5 to 1.7) | 0.2 (0.1 to 0.2) | 0.6 (0.5 to 0.7) | 1,593 (1,549 to 1,636) |
|  | 45 to 59y |  |  |  |  |  |
|  |  | January 1, 2022 to April 27, 2023 | 0.5 (0.4 to 0.5) | 0.2 (0.2 to 0.3) | 0.3 (0.2 to 0.3) | 1,341 (1,302 to 1,381) |
|  |  | April 28, 2023 to June 30, 2025 | 0.5 (0.5 to 0.6) | 0.1 (0.1 to 0.1) | 0.2 (0.2 to 0.3) | 1,553 (1,512 to 1,593) |
| Females |  |  |  |  |  |  |
|  | All |  |  |  |  |  |
|  |  | January 1, 2022 to April 27, 2023 | 6.0 (5.9 to 6.1) | 0.2 (0.1 to 0.2) | 0.8 (0.8 to 0.9) | 2,383 (2,357 to 2,409) |
|  |  | April 28, 2023 to June 30, 2025 | 5.5 (5.4 to 5.6) | 0.1 (0.1 to 0.1) | 0.6 (0.6 to 0.6) | 2,524 (2,435–2,628) |
|  | 16 to 24y |  |  |  |  |  |
|  |  | January 1, 2022 to April 27, 2023 | 19.9 (19.4 to 20.3) | 0.2 (0.1 to 0.2) | 2.3 (2.2 to 2.5) | 3,608 (3,535 to 3,680) |
|  |  | April 28, 2023 to June 30, 2025 | 18.0 (17.7 to 18.4) | 0.1 (0.1 to 0.1) | 1.7 (1.6 to 1.8) | 3,594 (3,530 to 3,658) |
|  | 25 to 34y |  |  |  |  |  |
|  |  | January 1, 2022 to April 27, 2023 | 6.7 (6.5 to 7.0) | 0.3 (0.2 to 0.3) | 1.0 (0.9 to 1.1) | 2,223 (2,175 to 2,271) |
|  |  | April 28, 2023 to June 30, 2025 | 6.5 (6.3 to 6.7) | 0.1 (0.1 to 0.2) | 0.8 (0.7 to 0.9) | 2,378 (2,335 to 2,422) |
|  | 35 to 44y |  |  |  |  |  |
|  |  | January 1, 2022 to April 27, 2023 | 2.1 (1.9 to 2.2) | 0.2 (0.1 to 0.2) | 0.4 (0.4 to 0.5) | 2,141 (2,091 to 2,192) |
|  |  | April 28, 2023 to June 30, 2025 | 1.9 (1.8 to 2.0) | 0.1 (0.1 to 0.1) | 0.3 (0.3 to 0.4) | 2,319 (2,275 to 2,365) |
|  | 45 to 59y |  |  |  |  |  |
|  |  | January 1, 2022 to April 27, 2023 | 0.5 (0.4 to 0.6) | 0.1 (0.1 to 0.1) | 0.1 (0.1 to 0.2) | 1,993 (1,950 to 2,036) |
|  |  | April 28, 2023 to June 30, 2025 | 0.5 (0.5 to 0.6) | 0.1 (0.0 to 0.1) | 0.1 (0.1 to 0.1) | 2,209 (2,168 to 2,251) |

CI: Confidence interval, estimated via bootstrap resampling with 10,000 iterations.
<sup>1</sup>We define chlamydia and gonorrhea cases as the first laboratory-confirmed detection of *C. trachomatis* and *N. gonorrhoeae*, respectively, at oral, rectal, or genital sites, in any 30-day period. We describe the algorithm for detection of incident syphilis cases in the supporting information (**Text S1**).
<sup>2</sup>One defined daily dose (DDD) of doxycycline is equivalent to 100 mg.
<sup>3</sup>We define at-risk MSM and TGW as those living with HIV or those with any history of receipt of HIV pre- or post-exposure prophylaxis from July 1, 2021 onward.

**Table 2:** Overall and indirect effects of doxyPEP among at-risk MSM and TGW.

| Outcome | Population | Incidence rate, cases per 1,000 person-years (95% CI) |  | Estimate (95% CI) |  |
|---|---|---|---|---|---|
|  |  | Observed | Expected<sup>1</sup> | Effect, %<sup>2</sup> | Incidence averted, cases per 1,000 person-years |
| Chlamydia |  |  |  |  |  |
|  | Overall effect | 80.2 (77.7 to 82.9) | 137.8 (120.2 to 158.6) | 41.8 (33.2 to 49.4) | 57.5 (39.9 to 78.3) |
|  | Indirect effect | 81.1 (78.5 to 83.8) | 136.2 (118.9 to 156.6) | 40.6 (31.9 to 48.3) | 55.1 (37.8 to 75.5) |
|  | Total effect | 36.7 (25.3 to 51.6) | 215.8 (171.8 to 266.5) | 80.0 (71.1 to 86.2) | 179.0 (135.1 to 229.8) |
| Syphilis |  |  |  |  |  |
|  | Overall effect | 21.3 (20.0 to 22.7) | 30.1 (24.6 to 37.1) | 29.2 (13.3 to 42.7) | 8.8 (3.3 to 15.8) |
|  | Indirect effect | 21.5 (20.2 to 22.9) | 30.0 (24.5 to 37.1) | 28.7 (12.5 to 42.1) | 8.5 (3.0 to 15.5) |
|  | Total effect | 10.0 (4.6 to 19.0) | 34.0 (21.1 to 51.1) | 64.8 (29.9 to 82.3) | 23.9 (11.1 to 41.1) |

CI: Confidence interval, estimated via bootstrap resampling with 10,000 iterations. We define at-risk MSM and TGW as those living with HIV or those with any history of receipt of HIV pre- or post-exposure prophylaxis from July 1, 2021 onward.
<sup>1</sup>We estimate expected incidence rates via interrupted time-series analyses excluding post-intervention changes in slopes, for incidence rates of each infection (**Figure 2**). Estimates among individuals who did not receive doxyPEP (indirect effects) exclude cases occurring within 90 days after doxyPEP dispenses.
<sup>2</sup>Estimated effects are reported as observed cases divided by expected cases.

These comparisons of incidence rates before and after doxyPEP implementation among all at-risk MSM and TGW correspond to “overall” effect estimates in field trials comparing risk among members of a population in which some individuals are exposed to an intervention versus a counterfactual scenario in which none are exposed to the intervention (**Figure 1**).^36,37^ Employing a test-negative design to distinguish direct effects of doxyPEP against each infection among at-risk recipients,^38^ we estimated that doxyPEP reduced incidence of chlamydia and syphilis among recipients by 66.3% (52.2 to 76.1%) and 50.1% (2.1 to 74.2%), respectively (**Table 3**). Corresponding analyses restricted to at-risk MSM and TGW meeting strict eligibility criteria for doxyPEP yielded similar point estimates (**Table S4**), as did analyses defining differing exposure periods connecting individuals’ dates of doxyPEP dispense and bSTI testing (**Tables S5**-**S6**). In contrast, we identified no evidence of direct protection against gonorrhea (–13.2% [–38.0 to 7.4%] risk reduction; **Table S7**).

**Table 3:** Direct effects of doxyPEP among at-risk MSM and TGW.

| **Outcome** | **DoxyPEP history within prior 90d, *n/N* (%)** |  | **Direct effect, % (95% CI)** |
|---|---|---|---|
|  | Cases (test positive) | Controls (test–negative) |  |
| Chlamydia | 33/3,705 (0.9) | 2,855/103,060 (2.8) | 66.3 (52.2 to 76.1) |
| Syphilis | 9/983 (0.9) | 2,732/99,982 (2.7) | 50.1 (2.1 to 74.2) |

CI: Confidence interval, estimated via bootstrap resampling with 10,000 iterations. We estimate direct effects via logistic regression models defining positive or negative test results as the outcome variable. Analyses include the first chlamydia test conducted in any 30-day period; we describe the algorithm for identifying incident syphilis infections in the supplementary appendix, which included >30-day restrictions on eligible tests for individuals with evidence of prior infection. We define direct effects as (1 − adjusted odds ratio) × 100%. We define at-risk MSM and TGW as those living with HIV or those with any history of receipt of HIV pre- or post-exposure prophylaxis from July 1, 2021 onward.

Indirect protection yielded 40.6% (31.9 to 48.3%) and 28.7% (12.5 to 42.1%) reductions in risk of chlamydia and syphilis, respectively, among non-recipients, without accompanying effects on gonorrhea incidence (0.0% [–14.0 to 11.9%] reduction; **Table 2**; **Figure 2**; **Table S3**). The substantial reduction in chlamydia and syphilis among non-recipients of doxyPEP—accounting for most cases prevented by the intervention (**Table S8**)—suggests doxyPEP implementation reached a core group of MSM and TGW wielding outsized influence on transmission of these infections among MSM and TGW more broadly. Among recipients, total effects of doxyPEP implementation corresponded to 80.0% (71.1 to 86.2%) and 64.8% (29.9 to 82.3%) reductions in risk of chlamydia and syphilis, respectively. These estimates remained similar in sensitivity analyses defining individuals’ doxyPEP history using a 180-day exposure window (**Table S9**).

### Dynamics of bSTIs among females and other males

To examine the role of MSM and TGW as a core group shaping bSTI transmission within the general population, we next tested whether the substantial reductions in chlamydia and syphilis incidence among MSM and TGW observed after doxyPEP implementation were further associated with indirect protection among females and other males. Females and heterosexual males were not eligible for doxyPEP under CDPH guidance implemented in 2023, and did not experience increases in doxycycline consumption commensurate with those occurring among at-risk MSM and TGW during the study period (**Figure 3**; **Tables S10**-**S11**). We therefore considered any change in incidence within these populations, relative to counterfactual expectations absent doxyPEP implementation, to represent indirect effects (**Figure 1**).

**Figure 3:**
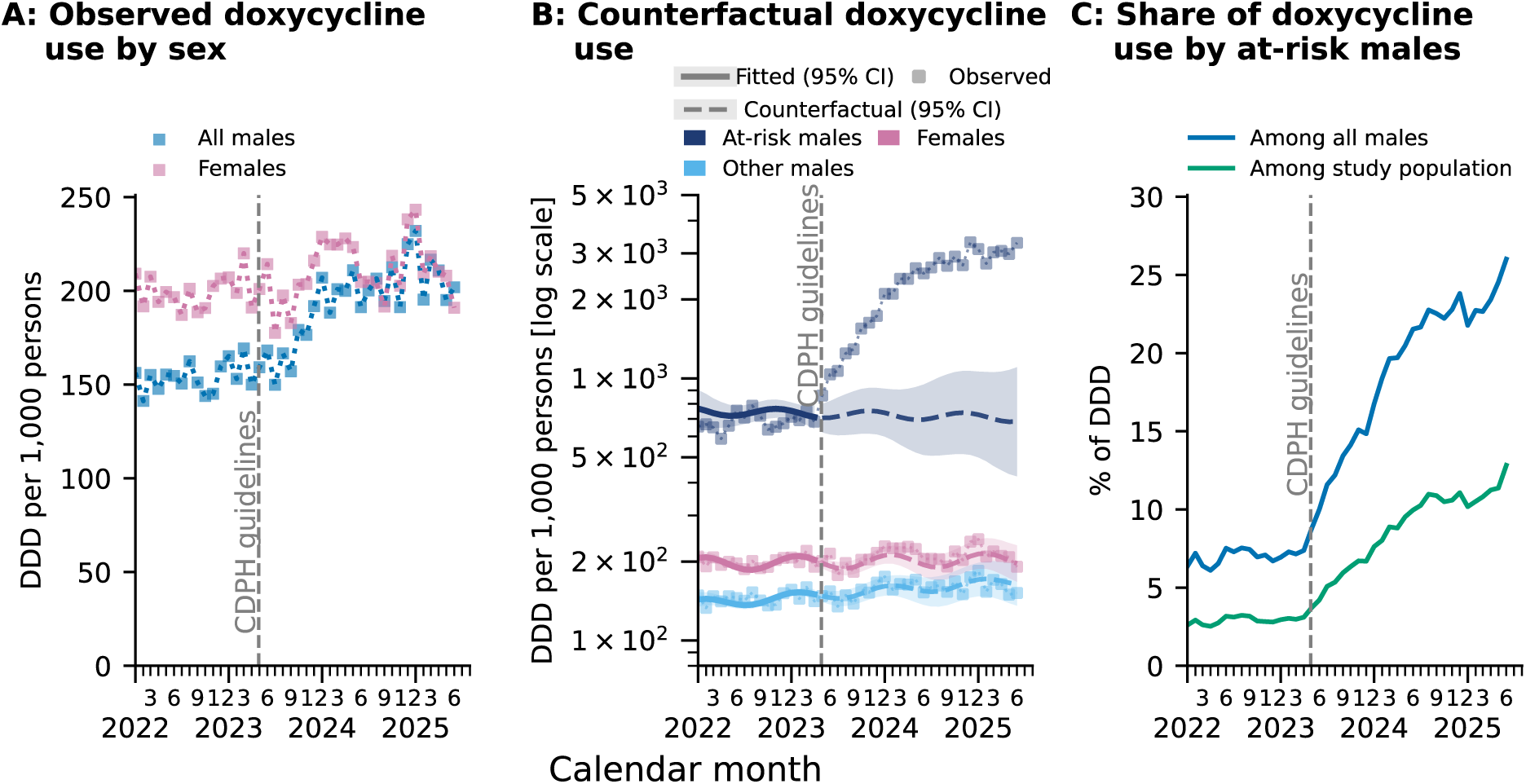
Doxycycline use within the study population. We illustrate (**a**) monthly doxycycline use, expressed as DDDs per 1,000 persons, among the full study cohort, stratified by sex, from January 2022 through June 2025. Next, we illustrate (**b**) observed and counterfactual monthly doxycycline use at-risk males (MSM/TGW living with HIV or with a history of HIV PrEP/PEP receipt), other males, and females. Counterfactual estimates were obtained using interrupted time-series models with time modeled as a continuous variable and adjusted for age group and seasonal variation. Dashed lines represent the counterfactual estimates, solid lines the fitted model, and points connected by dotted lines the observed data. Shaded areas delineate 95% confidence intervals. Last, we illustrate (**c**) the proportion of total doxycycline DDD use attributable to at-risk males among all males (blue line) and among the overall study cohort (green line). Vertical dashed lines indicate the publication date of the California doxyPEP guidelines. One doxycycline DDD was defined as 100 mg.

Incidence rates of chlamydia among females after doxyPEP implementation closely aligned with expectations projected from a continuation of pre-implementation trends (5.5 and 5.3 cases per 1,000 person-years respectively), consistent with observations for syphilis (0.1 cases per 1,000 person-years before and after implementation; **Table 4**; **Figure 2**). Incidence rates of chlamydia and syphilis were flat or increasing (–3.5% [–25.1 to 15.4%] and –4.8% [–70.0 to 37.9%] rate reductions, respectively), contrary to the declining trend among at-risk MSM and TGW; similar patterns were apparent in gonorrhea incidence. Among males not meeting the at-risk definition, incidence rates of chlamydia, syphilis, and gonorrhea closely resembled expectations from continuation of pre-implementation patterns (0.6% [–22.0 to 19.7%], 5.0% [–39.9 to 36.6%], and –0.7% [–32.1 to 23.4%] estimated reductions, respectively). These decoupled trends in bSTI incidence among at-risk MSM and TGW as compared to females and other males do not support the hypothesis that MSM and TGW act as a core group influencing bSTI transmission within broader sexual networks.

**Table 4:** Overall and indirect effects of doxyPEP among other males, and females.

| Population | Outcome | Population | Incidence rate, cases per 1,000 person-years (95% CI) |  | Estimate (95% CI) |  |
|---|---|---|---|---|---|---|
|  |  |  | Observed | Expected<sup>1</sup> | Effect, %<sup>2</sup> | Incidence averted, cases per 1,000 person-years |
| Other males<sup>3</sup> |  |  |  |  |  |  |
|  | Chlamydia | Without doxyPEP (indirect effect) | 2.8 (2.7 to 2.8) | 2.8 (2.3 to 3.5) | 0.6 (–22.0 to 19.7) | 0.0 (-–0.5 to 0.7) |
|  | Syphilis | Without doxyPEP (indirect effect) | 0.2 (0.2 to 0.2) | 0.2 (0.1 to 0.3) | 5.0 (–39.9 to 36.6) | 0.0 (–0.1 to 0.1) |
|  | Gonorrhea | Without doxyPEP (indirect effect) | 0.8 (0.8 to 0.9) | 0.8 (0.6 to 1.1) | –0.7 (–32.1 to 23.4) | 0.0 (–0.2 to 0.3) |
| Females |  |  |  |  |  |  |
|  | Chlamydia | Without doxyPEP (indirect effect) | 5.5 (5.4 to 5.6) | 5.3 (4.4 to 6.5) | –3.5 (–25.1 to 15.4) | –0.2 (–1.1 to 1.0) |
|  | Syphilis | Without doxyPEP (indirect effect) | 0.1 (0.1 to 0.1) | 0.1 (0.1 to 0.2) | –4.8 (–70.0 to 37.9) | 0.0 (-0.0 to 0.1) |
|  | Gonorrhea | Without doxyPEP (indirect effect) | 0.6 (0.6 to 0.6) | 0.5 (0.4 to 0.7) | –23.6 (–61.3 to 5.7) | –0.1 (–0.2 to 0.0) |

CI: Confidence interval, estimated via bootstrap resampling with 10,000 iterations.
<sup>1</sup>We estimate expected incidence rates via interrupted time-series analyses excluding post-intervention changes in slopes, for incidence rates of each infection (**Figure 2**). Estimates among individuals who did not receive doxyPEP (indirect effects) exclude cases occurring within 90 days after doxyPEP dispenses.
<sup>2</sup>Estimated effects are reported as observed cases divided by expected cases.
<sup>3</sup>Other males exclude at-risk MSM and TGW, defined as those living with HIV or those with any history of receipt of HIV pre- or post-exposure prophylaxis from July 1, 2021 onward.

### DoxyPEP uptake and doxycycline consumption

Core-group dynamics imply that preventive interventions may achieve substantial population impact even if allocated selectively to high-risk individuals ^39^. Such use strategies are a priority for doxyPEP, given the potential for negative externalities such as antimicrobial resistance selection in bSTIs as well as off-target pathogens^40,41^ potentially associated with broad population uptake. Although only 9% of at-risk males within the KPSC cohort received doxyPEP at any point during the study period, this increase in use corresponded to a 2.31-fold (1.99 to 2.64) rise in consumption, on average, among all at-risk MSM and TGW and 18,953 (16,052 to 21,048) additional defined daily doses of doxycycline per 1,000 at-risk MSM and TGW annually (**Figure 3**; **Table S10**-**S11**). Whereas at-risk MSM and TGW represented only 1.2% of individuals aged 16-59 years included in the study, their contribution to population-wide doxycycline consumption rose from 2.9% before doxyPEP implementation to 8.9% afterwards, and 12.8% by June, 2025.

## DISCUSSION

In southern California, direct protection against chlamydia and syphilis among MSM and TGW who initiated doxyPEP translated to onward reductions in cases among their counterparts who did not receive doxyPEP. This indirect effect suggests doxyPEP implementation reached a core group of high-risk MSM and TGW who played a prominent role sustaining bSTI transmission. The absence of a comparable indirect effect on gonorrhea—against which doxyPEP proved ineffective, in association with prevalent tetracycline resistance^27^—provides an implicit negative-control validation of our findings, as secular changes in bSTI testing or sexual behavior would be expected to drive similar trends in incidence rates of gonorrhea, chlamydia, and syphilis. However, reductions in bSTI incidence among at-risk MSM and TGW did not confer indirect protection to females and other males. This finding is inconsistent with the bisexual bridge theory positing that MSM and TGW, linked to heterosexual networks via MSMW, function as a core group influencing population-wide bSTI transmission.

Though supported by anecdotal examples of HIV and bSTI transmission to female partners of MSMW,^42^ the bisexual bridge theory has received scrutiny in several contexts. Genomic studies of *Neisseria gonorrhoeae* demonstrate that, although bridging occurs, distinct clusters of infection occur within MSM-and heterosexual networks,^10^ similar to observations in *Chlamydia trachomatis*^43,44^ and *Treponema pallidum* where lineages show clear associations with MSM and heterosexual case populations.^45–47^ Although illustrative of dynamics, these genomic studies provide little quantitative insight into the frequency or attributable role of bridging, and are limited by a lack of systematic sampling of cases. Epidemiologic studies encounter distinct challenges but have demonstrated lower bSTI prevalence among MSM with increasing proximity to women within their sexual networks,^48^ and lower bSTI risk factor prevalence among MSMW with greater numbers of female partners,^49,50^ undermining the notion that MSMW could play the central role implied by the bisexual bridge hypothesis.

Public health messaging surrounding doxyPEP implementation frequently has invoked the assumption of indirect effects extending to non-recipients including heterosexuals, in particularly noting rises in congenital syphilis infections as a premise motivating doxyPEP implementation among MSM and TGW.^51–53^ However, evidence of indirect effects of doxyPEP remains limited. Whereas several studies have reported overall reductions in chlamydia and syphilis incidence among MSM and TGW following doxyPEP implementation,^54,55^ a lack of individually-resolved antibiotic exposure data has made contributions of indirect protection to these trends unclear. In San Francisco, chlamydia incidence among cisgender women rose after doxyPEP implementation despite dramatic reductions among MSM and TGW,^54^ in line with our findings that trends in MSM and TGW do not drive trends within heterosexual networks. A separate study reported reductions in syphilis incidence among both cisgender males and females after doxyPEP implementation in King County, Washington,^56^ although findings merit scrutiny. Analyses projected a nearly 4-fold increase in syphilis incidence between 2020 and 2025 as a baseline expectation absent doxyPEP implementation, which may reflect pre-implementation changes in testing that do not generalize to other settings. In California, declines in syphilis incidence preceded doxyPEP implementation.^31^ The similar magnitude of estimated reductions in syphilis incidence among all males (52%) and females (47%) reported in the Washington study, despite doxyPEP implementation among MSM and TGW only, may likewise suggest a role of secular trends in explaining the study findings.

Strengths of our study include population representation across diverse racial, ethnic, and socioeconomic strata, with similar uptake of doxyPEP across groups; generalizability of KPSC members to the surrounding regional population;^57,58^ inclusion of >2 years of follow-up data after doxyPEP implementation; and well-characterized sexual healthcare utilization among individuals included in analyses of direct effects. However, there are several limitations. First, estimating indirect and overall effects requires assumptions that pre-implementation trends in bSTI incidence will continue after implementation. Nonetheless, biological plausibility of our findings is supported by their specificity: observed effects are restricted to chlamydia and syphilis, which remain susceptible to doxycycline, and to at-risk MSM and TGW, who are most likely to represent the sexual network of doxyPEP recipients. Second, unobserved differences among doxyPEP recipients and non-recipients in sexual risk or healthcare utilization may confound direct effects estimation. Although our analyses select on bSTI testing, differences in utilization of testing for screening or suspicion of STI symptoms cannot be measured. Last, receipt of doxyPEP through informal channels may not be tracked, and could result in exposure misclassification that would attenuate direct effect estimates and inflate indirect effect estimates among at-risk males. However, undocumented doxyPEP receipt is unlikely to be common in our study population due to availability of doxyPEP at no cost to KPSC members; moreover, eligibility for inclusion in our nested test-negative design study selects on utilization of KPSC for sexual health services, including HIV treatment or PrEP/PEP.

In our study, doxyPEP implementation among MSM and TGW at high risk of bSTIs provided an opportunity to probe the role of core groups in transmission. Indirect protection arising due to prevention of chlamydia and syphilis among doxyPEP recipients substantially lowered incidence of these infections among other MSM and TGW, suggesting the intervention reached individuals with a prominent role steering broader transmission within MSM and TGW networks. However, the lack of any discernable downstream reduction in cases among females and other males is inconsistent with the notion that MSM and TGW act as a core group influencing transmission within heterosexual networks. Our findings augment understanding of the public-health impact of doxyPEP, and should inform ongoing risk-benefit assessments addressing doxyPEP implementation strategies in differing populations.

## METHODS

### Study setting

The KPSC healthcare system provides integrated healthcare services across virtual, outpatient, emergency department, and inpatient settings to ∼5 million individuals throughout southern California. Members are enrolled through employer-sponsored, government-subsidized, and prepaid insurance schemes and broadly reflect the regional population with respect to demographic characteristics, socioeconomic status, and prevalence of common chronic conditions.^58^

### Design and study populations

We aimed to estimate overall, indirect, direct, and total of doxyPEP implementation on bSTI risk (**Figure 1**).^36^ Direct effects compared recipients’ risk against expectations under a counterfactual scenario where they did not receive doxyPEP. Indirect and overall effects compared risk against a counterfactual scenario without doxyPEP implementation, for non-recipients and for the full population (including recipients and non-recipients), respectively. Total effects compared recipients’ risk, as members of a population exposed to doxyPEP, against counterfactual risk under a scenario without doxyPEP implementation.

Our analyses considered three population groups of interest. We defined “at-risk” MSM and TGW as individuals assigned male sex at birth, aged 16-59 years, who were living with HIV or who had received HIV pre-exposure or post-exposure prophylaxis (PrEP/PEP; **Tables S12**-**S13**) any time since July 1, 2021. We separately analyzed “other” males (HIV-negative without PrEP/PEP history, most of whom were expected to have female partners) and females aged 16-59 years. We indexed follow-up time beginning on January 1, 2022 or 1 year after KPSC enrollment, whichever was later. We continued follow-up through death, disenrollment, or June 30, 2025, allowing for ≤45-day gaps in enrollment. Analyses included all individuals meeting eligibility criteria throughout their eligible follow-up periods.

### Outcomes

We defined gonorrhea and chlamydia infections as laboratory-confirmed detections of *N. gonorrhoeae* and *Chlamydia trachomatis*, respectively, by nucleic acid amplification tests of urethral, pharyngeal, or rectal specimens. We considered detections to represent incident infections if not preceded by any positive test for the same pathogen within 30 days. We defined incident syphilis using a longitudinal algorithm incorporating all available treponemal and non-treponemal test results and serial rapid plasma reagin (RPR) titers from individuals’ EHR (**Text S1**; **Figure S3**).

### Exposure

We defined doxyPEP prescription fills as outpatient doxycycline dispenses containing a ≥30-day supply, administered as 1×200mg tablets or 2×100mg tablets per dose, absent any laboratory-confirmed chlamydia or syphilis infection within 14 days before or after dispense. We quantified population-wide doxycycline use in defined daily doses (DDDs).^59^

### Estimands

We derived estimators for differences between observed and expected cases representing direct, indirect, overall, and total effects leveraging both time-series analyses of population incidence and case-control analyses of individual outcomes and time-varying doxyPEP exposure. We defined *Y*_0_ and *Y*_1_ as counts of observed cases among doxyPEP non-recipients and recipients, respectively, defined by history of any doxyPEP fill within 90 days before individuals’ date of testing. We let *Ŷ*_0_ and *Ŷ*_1_ indicate counts of expected cases, absent intervention, among non-recipients and recipients according to the definition above. We defined total expected cases, absent intervention, as *Ŷ*_0+1_=*Ŷ*_0_+*Ŷ*_1_. Thus, the difference *Ŷ*_0+1_-(*Ŷ*_0_+*Ŷ*_1_) conveyed cases prevented via overall effects; the difference *Ŷ*_0_-*Ŷ*_0_ conveyed cases prevented via indirect effects; and the difference *Ŷ*_1_-*Ŷ*_1_ conveyed cases prevented via total effects. We decomposed total effects among doxyPEP recipients to account separately for indirect and direct sources of protection. Defining *θ_I_* and *θ_D_* as risk ratios reflecting indirect and direct protection, respectively, yielded *Ŷ*_0_-θ_1_*Ŷ*_0_ and *Ŷ*_1_=θ_1_θ_D_*Ŷ*_1_, such that

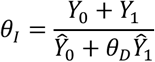

yielded the indirect effect when accounting for time-varying doxyPEP exposure.

### Identification strategy

#### Overall effects

We fit log-link negative binomial regression models to monthly case counts from January 1, 2022 to June 30, 2025 for each bSTI, specifying log-transformed person-time at risk as an offset in stratified analyses for at-risk MSM and TGW, other males (HIV-negative without history of PrEP/PEP receipt), and females. Models specified linear pre-and post-implementation slopes and intercepts, with adjustment for seasonality (**Text S1**). We estimated counterfactual incidence rates absent doxyPEP implementation by setting post-guideline slope changes to zero, thus projecting continuation of incidence according to the pre-implementation trend. We estimated cases averted due to overall effects among at-risk MSM and TGW via the difference between counterfactual and observed monthly case counts from May 1, 2023 to June 30, 2025, i.e. *Ŷ*_0+1_ − (*Ŷ*_0_ + *Ŷ*_1_), scaling incidence rate estimates to total (time-varying) population sizes. We additionally conducted probabilistic changepoint analyses to validate the temporal association of CDPH doxyPEP guidelines implementation with changes in slopes (**Figure S2**).^60^

#### Direct effects

We estimated direct effects of doxyPEP against laboratory-confirmed bSTIs among at-risk males via the test-negative design, aiming to recover the parameter *θ_D_* for each pathogen. We used logistic regression models to compute adjusted odds ratios (aORs) of doxyPEP dispense ≤90 days before specimen collection among individuals with positive versus negative laboratory test results for each bSTI between May 1, 2023 (after statewide guidance for doxyPEP implementation) and June 30, 2025, defining treatment effectiveness as (1 − aOR) × 100%. Under the test-negative design, the aOR recovers the risk ratio without rare disease assumptions^38^ (i.e., *θ_D_* = aOR). Models adjusted for prespecified confounders addressing demographic characteristics, healthcare utilization, indicators of sexual risk, and calendar time (**Text S1**; **Table S1**), and defined cluster-robust standard errors via the sandwich estimator to account for repeated testing among individuals. Restriction to individuals receiving bSTI testing was expected to mitigate bias from differences in bSTI testing frequency among doxyPEP recipients and non-recipients.

#### Indirect effects

We solved for the indirect effect parameter *θ_I_* among at-risk MSM and TGW via known and estimated quantities from the expression (*Y*_0_ + *Y*_1_)/(*Ŷ*_0_ + θ_D_*Ŷ*_1_). Here, *Y*_0_ and *Y*_1_ were known from KPSC data, and *θ_D_* was estimable as the direct effect via the test-negative design, leaving *Ŷ*_0_ and *Ŷ*_1_ unknown. Our overall effects analysis projected *Ŷ*_0+1_ from pre-implementation trends in incidence of each bSTI among all individuals. To partition this sum across a dynamic population of doxyPEP recipients and non-recipients, we sampled from the distribution of monthly estimates for *Ŷ*_0+1_ and assigned the sampled cases, at random, to individuals within the at-risk MSM and TGW cohort, independent of doxyPEP receipt history while accounting for other measured predictors of individuals’ likelihood of receiving bSTI diagnoses. We sampled individuals at random using propensity weights from logistic regression models defining each bSTI as an outcome variable and all measured covariates as predictors (**Text S1**). We fit propensity weights models to data from June 2022 to April 2023 to exclude differences relating to doxyPEP implementation. We defined *Ŷ*_1_ during each month as the sum of predicted cases assigned to individuals who had received doxyPEP in the preceding 90 days, and *Ŷ*_0_ as the sum of predicted cases assigned to individuals who had not received doxyPEP in the preceding 90 days. In sensitivity analyses, we repeated this procedure using a 180-day doxyPEP exposure window. We input values from 10,000 draws of *Ŷ*_0_, *Ŷ*_1_, and *θ_D_* to reconstruct the distribution of *θ_I_*.

As most males and females outside the at-risk population were unlikely to be MSM or TGW, and thus ineligible for doxyPEP, we considered differences between estimated counterfactual and observed case counts in these populations to represent indirect effects.

### Ethics

The Kaiser Permanente Southern California Institutional Review Board reviewed and approved the study.

## Data Availability

The raw data used in this manuscript include protected health information (e.g., dates of diagnoses and testing) that cannot be shared openly without appropriate ethical oversight and approval as well as institutional data use agreements. Kaiser Permanente Southern California (KPSC) institutional policy requires a data transfer agreement to be executed between KPSC and the individual recipient entity prior to transmittal of patient-level data outside KPSC. Requests for data can be addressed to kpsc.irb{at}kp.org.

## FUNDING

This work was funded by the US Centers for Disease Control & Prevention (CDC--RFA-FT-23-0069 to SYT and JAL).

## AUTHOR CONTRIBUTIONS

MY, SYT, and JAL conceptualized the study; MY, JTL, and JAL devised the methodology; MY, KP, VH, and GSD conducted the investigation; MY generated figures; JAL and SYT acquired funding; IACR and MEP provided project administration; IRB, NFM, SYT, and JAL provided supervision; MY and JAL wrote the first draft of the manuscript; BK, VH, JTL, IACR, MEP, GSD, IRB, NFM, and SYT contributed to manuscript review and editing.

## COMPETING INTERESTS STATEMENT

The authors declare no competing interests.

## DATA AVAILABILITY

Individual-level testing and clinical outcomes data reported in this study are not publicly shared due to privacy protections for patient electronic health records. Individuals wishing to access disaggregated data, including data reported in this study, should submit requests for access to. Requests will receive a response within 14 days. De-identified data (including, as applicable, participant data and relevant data dictionaries) will be shared upon approval of analysis proposals with signed data-access agreements in place.

## CODE AVAILABILITY

Analysis code is publicly available without restriction from the corresponding author’s “doxyPepIndirects” GitHub repository (joelewnard/doxyPepIndirects).

### Text S1: Expanded methods

#### 1: Study population

We analyzed EHRs of all KPSC members aged 16-59 years from three population groups. We defined “at-risk” males as individuals assigned male at birth, born on or before 30 September, 2008, and who were living with HIV infection or receiving emtricitabine/tenofovir disoproxil fumarate (oral) or emtricitabine/tenofovir alafenamide (injectable) between 1 July, 2021, and 30 June, 2025. We classified members as living with HIV infection if listed in the HIV Registry as of June 30, 2025, or if they had at least 3 encounters with any of the specified HIV diagnosis codes since 2015 (**Table S11**); classification as living with HIV or receiving HIV pre-or post-exposure prophylaxis began on the first date these definitions were satisfied.

We considered other males (HIV negative without PrEP history) and females unlikely to be eligible for doxyPEP under current guidelines and analyzed them as separate groups.

Study follow-up commenced from the latest of the following dates: 1 January, 2022, one year after the date of health plan entry, or members’ 16^th^ birthday. Follow-up continued through the earliest of: 30 June, 2025, death, or disenrollment (allowing for lapses in membership lasting up to 45 days).

#### 2: Syphilis testing algorithm

We used longitudinal syphilis laboratory testing data collected between July 2017 and June 2025 to identify incident syphilis infections within the study population (**Figure S3**). Laboratory data included treponemal assays (enzyme immunoassay [EIA], *Treponema pallidum* particle agglutination assay [TPPA], and agglutination assays [AGGL]) and quantitative non-treponemal rapid plasma reagin (RPR) titers. When multiple laboratory results of the same test type were available for a given specimen collection date, results were rolled into a single record. For RPR testing, the highest reported titer was retained, consistent with CDC laboratory recommendations.^1^ When multiple treponemal test results were available from the same collection date, any reactive result was considered evidence of treponemal reactivity. This approach preserves evidence of prior treponemal reactivity in the longitudinal serologic history, preventing individuals with discordant historical results from being incorrectly classified as having no prior infection. Classifying discordant results as nonreactive increased incident case ascertainment by only 33 cases (<0.3%), with no meaningful impact on study findings.

We adapted our longitudinal algorithm for distinguishing incident syphilis infections from previously published approaches.^2,3^ Among individuals without prior laboratory testing (**Figure S3a**), cases identified by RPR testing alone were classified as incident syphilis only when the RPR titer was ≥1:16. When both treponemal and non-treponemal testing were available, incident syphilis required a reactive treponemal test and an RPR titer ≥1:8. We selected these thresholds to reduce false-positive case classification resulting from left censoring of historical laboratory data and the absence of diagnostic coding, in line with previous approaches.

For individuals with prior laboratory testing, we evaluated changes in RPR titers longitudinally. We defined a significant increase as a ≥4-fold rise from the reference RPR titer or,^4^ when the reference RPR result was nonreactive, a current RPR titer ≥1:4.

Among individuals with prior laboratory testing but no previously identified syphilis episode (**Figure S3b**), we classified incident infection differentially according to recent serologic history. If no prior RPR result was available within the preceding 24 months, incident syphilis required a reactive treponemal test and an RPR titer ≥1:8. Otherwise, incident infection required a ≥4-fold increase in RPR titer compared with the most recently-obtained RPR result within the previous 24 months. We defined the 24-month window to align with recommendations for interpreting serologic response following syphilis treatment, particularly among individuals living with HIV.^4^

Among individuals with a previously identified syphilis episode, we defined recurrent infection based on a ≥4-fold increase in RPR titer from the post-treatment nadir following the most recent infection, consistent with current syphilis management recommendations.^4^ We classified observations as recurrent infections when the ≥4-fold increase occurred at least 365 days after the previous episode. If the increase occurred between 180 and 364 days after the previous episode, we required an RPR titer ≥1:8 to improve specificity; otherwise, no new infection was assigned. For individuals living with HIV at the time of testing, these intervals were extended to 730 and 365 days, respectively, to account for the slower and more variable decline in non-treponemal titers observed after treatment in this population.^4,5^

#### 3: Direct effects estimation

Models included data from testing among at-risk MSM and TGW, and adjusted for prespecified confounders, including age at testing (continuous); race or ethnicity (Hispanic (any race), White (non-Hispanic), Black (non-Hispanic), Asian/Pacific Islander (non-Hispanic), Other/Unknown); cisgender or transgender identity (ascertained using natural language processing of structured and unstructured EHR data;^6^ binary); HIV infection status or receipt of HIV PrEP/PEP during the preceding six months (“0”, HIV infection, HIV PrEP/PEP); history of STI testing and STI diagnoses during the preceding six months (continuous), prior JYNNEOS vaccination (binary); commercial or non-commercial source of insurance coverage (binary); community-level socioeconomic status, as measured by neighborhood deprivation index (continuous); calendar month (continuous); and seasonal trends (sine and cosine transformation of calendar date with 12-month periodicity). We defined cluster-robust standard errors to account for repeated testing within individuals. Sensitivity analyses considered direct effects by bSTI history (**Table S4**), by period (2023, 2024, and 2025 calendar years; **Table S5**) and with doxyPEP exposure history defined by any prescription dispense within 180 days before testing (**Table S6**).

#### 4: Overall effects estimation

We estimated the overall effect of doxyPEP implementation on bSTIs and doxycycline use among at-risk MSM and TGW, other males, and females using interrupted time-series (ITS) analyses with negative binomial regression models specifying a log link. We specified outcomes as monthly counts of laboratory-confirmed cases for each bSTI, by population stratum; models included terms for pre-and post-guideline intercepts and slopes, age group (<25y, 25-34y, 44-59y), and log-transformed person-time at risk as an offset; we modeled calendar time as a continuous variable, and adjusted for seasonal variation using sine and cosine transformations of calendar time (calculated at midpoints of each month) with 12-month periodicity.

We modeled counterfactual incidence rates and case counts using the fitted models assuming no doxyPEP implementation, i.e., setting the post-guideline slope changes to zero while preserving the estimated pre-guideline trend. Fitted models estimated a statistically-significant, positive intercept at the time of doxyPEP implementation among at-risk MSM and TGW for all bSTIs (**Figure 2**), likely attributable to an increase in testing driven by individuals who received doxyPEP (**Figure S4**). We thus accounted for the post-implementation change in intercept when estimating counterfactual incidence among at-risk MSM and TGW to control for contributions of this increase in testing to rates of bSTI detection. As similar changes in intercept did not appear among females or other males, primary analyses for these population strata considered changes in slopes only after doxyPEP implementation (**Table 3**; **Figure 2**). Sensitivity analyses further allowing for a change in intercept at the time of doxyPEP implementation among females and other males yielded similar findings (**Table S14**), as did analyses assuming shared slopes in bSTI incidence rates among females and other males (**Table S15**; **Figure S5**-**S6**).

To determine whether incidence time series supported changes in slopes around the time of statewide doxyPEP implementation (April 28, 2023), we fit changepoint models assuming either (*a*) no change in slope or (*b*) models defining changes in slope at each candidate month from July 2022 to December 2024, leaving a 6-month period at both the beginning and end of the study period to mitigate edge effects. We compared the fit of these models via Bayesian Information Criterion (BIC) scores. We illustrate outputs via averaged month-specific slopes, based on BIC weights across all fitted models (**Figure S2**).

#### 5: Indirect effects estimation

We used a model-based approach to estimate indirect effects among at-risk MSM and TGW, defining

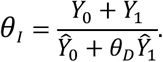

where *θ_I_* was the rate ratio relating observed to expected incidence among untreated individuals, such that the indirect effect was equal to (1 − *θ_I_*) × 100%; *Ŷ*_0_ and *Ŷ*_1_ were observed case counts without and with doxyPEP dispenses in the preceding 90 days, respectively; *Ŷ*_0_ and *Ŷ*_1_ were expected case counts among the same population groups absent doxyPEP implementation; and *θ_D_* = *aOR* conveyed the direct effect estimate from our test-negative design analyses.

We sampled the sum *Ŷ*_0+1_ = *Ŷ*_0_+*Ŷ*_1_, by month, from projected incidence rates absent doxyPEP implementation (see section 4: “Overall effects estimation”, above) and partitioned cases among doxyPEP recipients and non-recipients at random to determine *Ŷ*_0_ and *Ŷ*_1_ To determine individuals’ relative propensity of each bSTI, absent doxyPEP receipt, we fit logistic regression models defining all variables included in our adjustment set for direct effects (see section 3, “Direct effects estimation”, above) as predictors, with an outcome of any diagnosis of each bSTI over the pre-implementation period of June 2022 to April 2023. We assigned predicted cases to individuals based on the resulting propensity weights, ignoring doxyPEP receipt, and defined *Ŷ*_0_ as the sum of predicted cases assigned to individuals who had not received doxyPEP in the preceding 90 days and *Ŷ*_1_ as the sum of predicted cases assigned to individuals who had received doxyPEP in the preceding 90 days. We drew 10,000 draws from the monthly sums *Ŷ*_0+1_ and generated corresponding *Ŷ*_0_ and *Ŷ*_1_ values at random for each draw. We generated 10,000 independent draws from the distribution of *θ_D_*.

#### 6: Estimation of effects on doxycycline consumption

We fit ITS models as described above for overall effects defining monthly DDDs of doxycycline dispensed as the outcome variable. We defined counterfactual rates setting both post-guidelines slopes and intercepts to zero, as changes in intercept were expected to reflect new doxyPEP initiation or treatment of infections identified as a consequence of enhanced testing undertaken due to doxyPEP initiation.

---

**Table S1:**
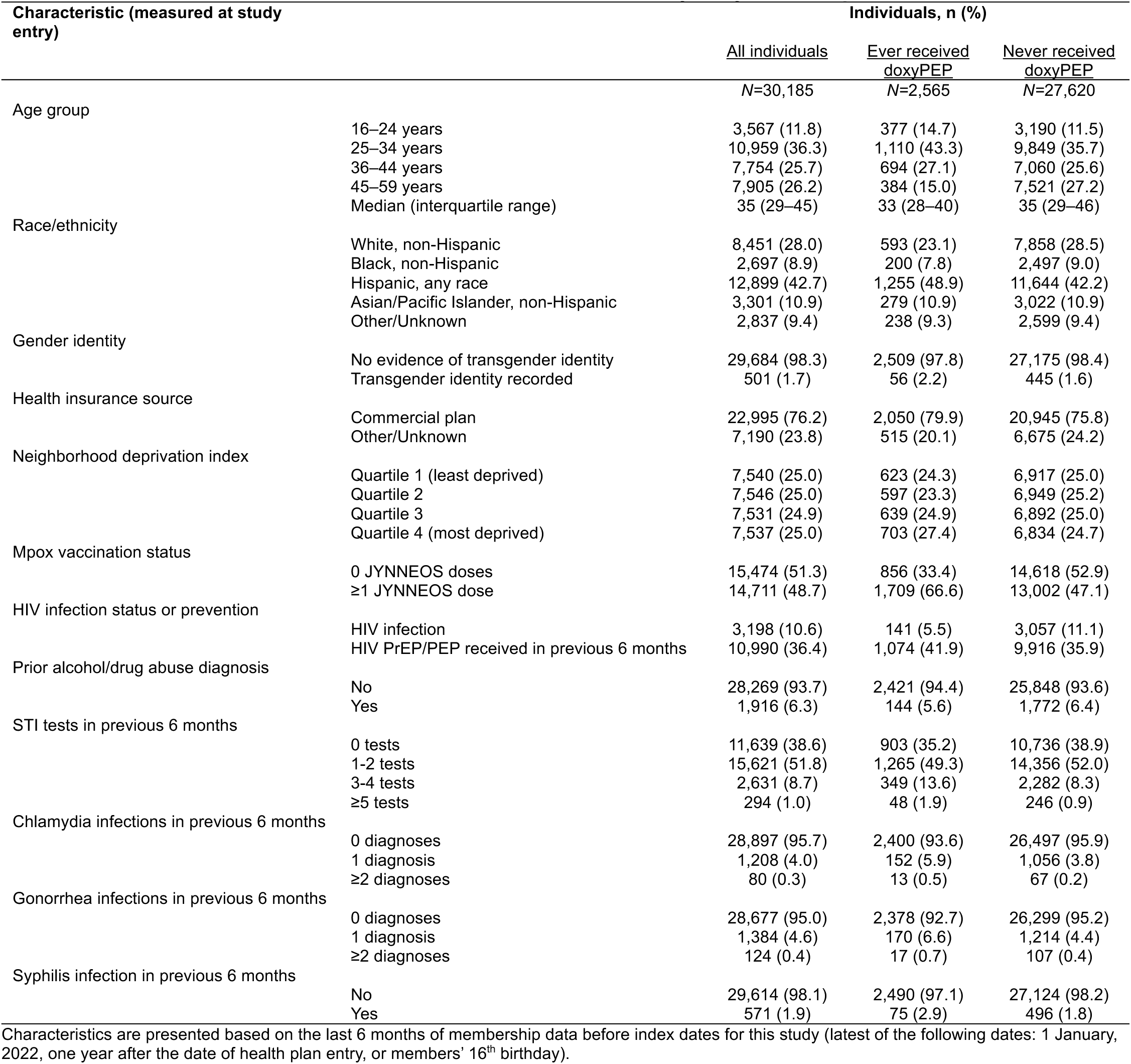
Baseline characteristics of at-risk MSM and TGW cohort, by doxyPEP receipt.

| **Characteristic (measured at study entry)** |  | **Individuals, n (%)** |  |  |
|---|---|---|---|---|
|  |  | All individuals | Ever received doxyPEP | Never received doxyPEP |
|  |  | *N*=30,185 | *N*=2,565 | *N*=27,620 |
| Age group |  |  |  |  |
|  | 16–24 years | 3,567 (11.8) | 377 (14.7) | 3,190 (11.5) |
|  | 25–34 years | 10,959 (36.3) | 1,110 (43.3) | 9,849 (35.7) |
|  | 36–44 years | 7,754 (25.7) | 694 (27.1) | 7,060 (25.6) |
|  | 45–59 years | 7,905 (26.2) | 384 (15.0) | 7,521 (27.2) |
|  | Median (interquartile range) | 35 (29–45) | 33 (28–40) | 35 (29–46) |
| Race/ethnicity |  |  |  |  |
|  | White, non-Hispanic | 8,451 (28.0) | 593 (23.1) | 7,858 (28.5) |
|  | Black, non-Hispanic | 2,697 (8.9) | 200 (7.8) | 2,497 (9.0) |
|  | Hispanic, any race | 12,899 (42.7) | 1,255 (48.9) | 11,644 (42.2) |
|  | Asian/Pacific Islander, non-Hispanic | 3,301 (10.9) | 279 (10.9) | 3,022 (10.9) |
|  | Other/Unknown | 2,837 (9.4) | 238 (9.3) | 2,599 (9.4) |
| Gender identity |  |  |  |  |
|  | No evidence of transgender identity | 29,684 (98.3) | 2,509 (97.8) | 27,175 (98.4) |
|  | Transgender identity recorded | 501 (1.7) | 56 (2.2) | 445 (1.6) |
| Health insurance source |  |  |  |  |
|  | Commercial plan | 22,995 (76.2) | 2,050 (79.9) | 20,945 (75.8) |
|  | Other/Unknown | 7,190 (23.8) | 515 (20.1) | 6,675 (24.2) |
| Neighborhood deprivation index |  |  |  |  |
|  | Quartile 1 (least deprived) | 7,540 (25.0) | 623 (24.3) | 6,917 (25.0) |
|  | Quartile 2 | 7,546 (25.0) | 597 (23.3) | 6,949 (25.2) |
|  | Quartile 3 | 7,531 (24.9) | 639 (24.9) | 6,892 (25.0) |
|  | Quartile 4 (most deprived) | 7,537 (25.0) | 703 (27.4) | 6,834 (24.7) |
| Mpox vaccination status |  |  |  |  |
|  | 0 JYNNEOS doses | 15,474 (51.3) | 856 (33.4) | 14,618 (52.9) |
|  | ≥1 JYNNEOS dose | 14,711 (48.7) | 1,709 (66.6) | 13,002 (47.1) |
| HIV infection status or prevention |  |  |  |  |
|  | HIV infection | 3,198 (10.6) | 141 (5.5) | 3,057 (11.1) |
|  | HIV PrEP/PEP received in previous 6 months | 10,990 (36.4) | 1,074 (41.9) | 9,916 (35.9) |
| Prior alcohol/drug abuse diagnosis |  |  |  |  |
|  | No | 28,269 (93.7) | 2,421 (94.4) | 25,848 (93.6) |
|  | Yes | 1,916 (6.3) | 144 (5.6) | 1,772 (6.4) |
| STI tests in previous 6 months |  |  |  |  |
|  | 0 tests | 11,639 (38.6) | 903 (35.2) | 10,736 (38.9) |
|  | 1-2 tests | 15,621 (51.8) | 1,265 (49.3) | 14,356 (52.0) |
|  | 3-4 tests | 2,631 (8.7) | 349 (13.6) | 2,282 (8.3) |
|  | ≥5 tests | 294 (1.0) | 48 (1.9) | 246 (0.9) |
| Chlamydia infections in previous 6 months |  |  |  |  |
|  | 0 diagnoses | 28,897 (95.7) | 2,400 (93.6) | 26,497 (95.9) |
|  | 1 diagnosis | 1,208 (4.0) | 152 (5.9) | 1,056 (3.8) |
|  | ≥2 diagnoses | 80 (0.3) | 13 (0.5) | 67 (0.2) |
| Gonorrhea infections in previous 6 months |  |  |  |  |
|  | 0 diagnoses | 28,677 (95.0) | 2,378 (92.7) | 26,299 (95.2) |
|  | 1 diagnosis | 1,384 (4.6) | 170 (6.6) | 1,214 (4.4) |
|  | ≥2 diagnoses | 124 (0.4) | 17 (0.7) | 107 (0.4) |
| Syphilis infection in previous 6 months |  |  |  |  |
|  | No | 29,614 (98.1) | 2,490 (97.1) | 27,124 (98.2) |
|  | Yes | 571 (1.9) | 75 (2.9) | 496 (1.8) |

Characteristics are presented based on the last 6 months of membership data before index dates for this study (latest of the following dates: 1 January, 2022, one year after the date of health plan entry, or members' 16<sup>th</sup> birthday).

**Table S2:** Distribution of doxyPEP prescription fills, by dose and quantity dispensed.

| **Doxycycline dose** | **Quantity dispensed, tablets** | **Prescriptions, *n* (%)** |
|---|---|---|
|  |  | *N*=5,530 |
| 100 mg |  |  |
|  | 60 | 5,076 (91.8) |
|  | 90 | 157 (2.8) |
|  | 120 | 153 (2.8) |
|  | 180 | 136 (2.5) |
| 200 mg |  |  |
|  | 30 | 8 (0.1) |
| Mean DDDs dispensed (95% CI) |  |  |
|  | 65.5 (64.9 to 66.0) |  |

**Table S3:** Overall, indirect, and total effects of doxyPEP on gonorrhea among at-risk MSM and TGW.

| **Population** | **Incidence rate, cases per 1,000 person-years (95% CI)** |  | **Estimate (95% CI)** |  |
|---|---|---|---|---|
|  | Observed | Expected<sup>1</sup> | Effect, %<sup>2</sup> | Incidence averted, cases per 1,000 person-years |
| Overall effect | 121.8 (118.6 to 125.0) | 120.8 (106.2 to 137.3) | –0.9 (–14.7 to 11.3) | –1.0 (–15.7 to 15.5) |
| Indirect effect | 119.8 (116.6 to 123.0) | 119.4 (105.0 to 135.9) | 0.0 (–14.0 to 11.9) | –0.4 (–14.8 to 16.1) |
| Total effect | 224.8 (194.8 to 258.0) | 191.0 (152.9 to 236.0) | –21.9 (–48.8 to –0.6) | –33.7 (–71.8 to 11.2) |

CI: Confidence interval, estimated via bootstrap resampling with 10,000 iterations. We define at-risk MSM and TGW as those living with HIV or those with any history of receipt of HIV pre- or post-exposure prophylaxis from July 1, 2021 onward.
<sup>1</sup>We estimate expected incidence rates via interrupted time-series analyses excluding post-intervention changes in slopes, for incidence rates of each infection (**Figure 2**). Estimates among individuals who did not receive doxyPEP (indirect effects) exclude cases occurring within 90 days after doxyPEP dispenses.
<sup>2</sup>Estimated effects are reported as observed cases divided by expected cases.

**Table S4:** Direct effects of doxyPEP within analyses restricted to at-risk MSM and TGW with bSTI diagnoses within 6 or 12 months before testing.

| Outcome | Population stratum | DoxyPEP history within prior 90d, *n/N* (%) |  | Direct effect, % (95% CI) |
|---|---|---|---|---|
|  |  | Cases (test positive) | Controls (test–negative) |  |
| Chlamydia |  |  |  |  |
|  | As recommended (STI in prior 12 months) | 19/1,601 (1.2) | 1,111/25,402 (4.4) | 66.8 (47.6 to 78.7) |
|  | Stringent criteria (STI in prior 6 months) | 12/932 (1.3) | 671/14,253 (4.7) | 65.9 (39.6 to 80.6) |
| Syphilis |  |  |  |  |
|  | As recommended (STI in prior 12 months) | 5/269 (1.9) | 1,017/24,470 (4.2) | 46.0 (–33.0 to 78.3) |
|  | Stringent criteria (STI in prior 6 months) | 1/111 (0.9) | 596/13,355 (4.5) | 75.3 (-84.2 to 96.5) |

CI: Confidence interval, estimated via bootstrap resampling with 10,000 iterations. We define at-risk MSM and TGW as those living with HIV or those with any history of receipt of HIV pre- or post-exposure prophylaxis from July 1, 2021 onward. We estimate direct effects via logistic regression models defining positive or negative test results as the outcome variable. Analyses include the first chlamydia test conducted in any 30-day period; we describe the algorithm for identifying incident syphilis infections in the supplementary appendix, which included >30-day restrictions on eligible tests for individuals with evidence of prior infection. We define direct effects as (1 − *adjusted odds ratio*) × 100%.

**Table S5:** Direct effects of doxyPEP among at-risk MSM and TGW, by period.

| Period | Outcome | **DoxyPEP history within prior 90d, *n/N* (%)** |  | **Estimate (95% CI)** |  |
|---|---|---|---|---|---|
|  |  | Cases (test positive) | Controls (test–negative) | Direct effect, % | Incidence averted, cases per 1000 person–years |
| May-December, 2023 |  |  |  |  |  |
|  | Chlamydia | 13/1,684 (0.8) | 337/30,702 (1.1) | 43.8 (0.5 to 68.9) | 83.0 (0.5 to 264.9) |
|  | Syphilis | 1/447 (0.2) | 326/30,354 (1.1) | 78.6 (–59.8 to 97.0) | 30.1 (–3.4 to 477.6) |
|  | Gonorrhea | 6/1,885 (1.4) | 324/30,502 (1.1) | 1.8 (–48.9 to 35.6) | 3.8 (–74.9 to 122.9) |
| January-December, 2024 |  |  |  |  |  |
|  | Chlamydia | 13/1,496 (0.9) | 1,493/48,049 (3.1) | 73.4 (54.4 to 84.5) | 77.8 (27.7 to 184.7) |
|  | Syphilis | 6/406 (1.5) | 1,424/46,523 (3.1) | 33.2 (–48.1 to 70.8) | 6.5 (–4.6 to 37.3) |
|  | Gonorrhea | 108/2,585 (4.2) | 1,398/46,959 (3.0) | –19.8 (–47.9 to 2.7) | –38.6 (–77.2 to 6.7) |
| January-June, 2025 |  |  |  |  |  |
|  | Chlamydia | 7/525 (1.3) | 1,025/24,309 (4.2) | 68.4 (32.0 to 85.1) | 48.6 (8.9 to 161.1) |
|  | Syphilis | 2/130 (1.5) | 982/23,105 (4.3) | 42.3 (–136.2 to 85.7) | 4.7 (–5.1 to 57.4) |
|  | Gonorrhea | 68/1,154 (5.9) | 962/23,680 (4.1) | –36.3 (–77.3 to –3.8) | –57.9 (–101.3 to –8.0) |

CI: Confidence interval, estimated via bootstrap resampling with 10,000 iterations. We define at-risk MSM and TGW as those living with HIV or those with any history of receipt of HIV pre- or post-exposure prophylaxis from July 1, 2021 onward. We estimate direct effects via logistic regression models defining positive or negative test results from the first test conducted in any 30-day period as the outcome variable. We define direct effects as (1 − *adjusted odds ratio*) × 100%.

**Table S6:** Direct effects of doxyPEP among at-risk MSM and TGW, applying a 180-day exposure history definition.

| **Outcome** | **Population stratum** | **DoxyPEP history within prior 180d, *n/N* (%)** |  | **Estimate (95% CI)** |  |
|---|---|---|---|---|---|
|  |  | Cases (test positive) | Controls (test–negative) | Direct effect, % | Incidence averted, cases per 1000 person–years |
| Chlamydia |  |  |  |  |  |
|  | As recommended (STI in prior 12 months) | 35/1,601 (2.2) | 1,739/25,402 (6.8) | 61.0 (45.5 to 72.0) | 101.2 (50.0 to 182.8) |
|  | As implemented (all at-risk<sup>1</sup>) | 64/3,705 (1.7) | 4,550/103,060 (4.4) | 58.7 (47.1 to 67.8) | 65.4 (38.5 to 102.8) |
|  | Stringent criteria (STI in prior 6 months) | 21/932 (2.3) | 1,019/14,253 (7.1) | 60.6 (39.4 to 74.2) | 107.0 (42.3 to 221.0) |
| Syphilis |  |  |  |  |  |
|  | As recommended (STI in prior 12 months) | 7/269 (2.6) | 1,613/24,470 (6.6) | 53.1 (-0.6 to 78.3) | 15.5 (–0.1 to 61.8) |
|  | As implemented (all at-risk<sup>1</sup>) | 12/983 (1.2) | 4,382/99,982 (4.4) | 58.9 (26.5 to 76.8) | 12.4 (2.8 to 33.9) |
|  | Stringent criteria (STI in prior 6 months) | 2/111 (1.8) | 918/13,355 (6.9) | 67.7 (-33.8 to 92.1) | 18.2 (–2.2 to 139.6) |

CI: Confidence interval, estimated via bootstrap resampling with 10,000 iterations. We define at-risk MSM and TGW as those living with HIV or those with any history of receipt of HIV pre- or post-exposure prophylaxis from July 1, 2021 onward. We estimate direct effects via logistic regression models defining positive or negative test results from the first test conducted in any 30-day period as the outcome variable. We define direct effects as (1 − *adjusted odds ratio*) × 100%.

**Table S7:** Direct effect of doxyPEP against gonorrhea among at-risk MSM and TGW.

| **Population stratum** | **DoxyPEP history within prior 90d, *n/N* (%)** |  | **Estimate (95% CI)** |  |
|---|---|---|---|---|
|  | Cases (test positive) | Controls (test–negative) | Direct effect, % | Incidence averted, cases per 1000 person–years |
| As-recommended (STI in prior 12 months) | 113/2,520 (4.5) | 1,016/24,480 (4.2) | –13.2 (–38.0 to 7.4) | –42.9 (–104.5 to 29.6) |
| As implemented (all at-risk) | 202/5,624 (3.6) | 2,684/101,141 (2.7) | –21.9 (–41.5 to –4.7) | –40.3 (–67.4 to –10.0) |
| Stringent criteria (STI in prior 6 months) | 62/1,471 (4.2) | 620/13,710 (4.5) | 1.3 (–29.6 to 24.8) | 4.6 (–82.3 to 121.1) |

CI: Confidence interval, estimated via bootstrap resampling with 10,000 iterations. We define at-risk MSM and TGW as those living with HIV or those with any history of receipt of HIV pre- or post-exposure prophylaxis from July 1, 2021 onward. We estimate direct effects via logistic regression models defining positive or negative test results from the first test conducted in any 30-day period as the outcome variable. We define direct effects as $(1 - adjusted\ odds\ ratio) \times 100\%$.

**Table S8:** Estimated chlamydia, syphilis, and gonorrhea cases averted following doxyPEP implementation.

| Population | Outcome | DoxyPEP exposure group | Cases, *n* (95% CI) |  |  |
|---|---|---|---|---|---|
|  |  |  | Expected | Observed | Averted |
| At-risk MSM and TGW<sup>1</sup> |  |  |  |  |  |
|  | Chlamydia |  |  |  |  |
|  |  | All (overall effect) | 6,361 (5,548 to 7,321) | 3,705 (3,587 to 3,826) | 2,656 (1,843 to 3,616) |
|  |  | Non-recipients (indirect effect) | 6,167 (5,384 to 7,090) | 3,672 (3,554 to 3,793) | 2,495 (1,712 to 3,418) |
|  |  | Recipients (direct effect) | 194 (154 to 240) | 33 (23 to 46) | 161 (121 to 207) |
|  | Syphilis |  |  |  |  |
|  |  | All (overall effect) | 1,389 (1,134 to 1,714) | 983 (923 to 1,046) | 406 (151 to 731) |
|  |  | Non-recipients (indirect effect) | 1359 (1108 to 1678) | 974 (914 to 1037) | 385 (134 to 704) |
|  |  | Recipients (direct effect) | 31 (19 to 46) | 9 (4 to 17) | 22 (10 to 37) |
|  | Gonorrhea |  |  |  |  |
|  |  | All (overall effect) | 5,576 (4,901 to 6,341) | 5,624 (5,478 to 5,773) | -48 (-723 to 717) |
|  |  | Non-recipients (indirect effect) | 5,404 (4753 to 6151) | 5,422 (5279 to 5568) | -18 (-669 to 729) |
|  |  | Recipients (direct effect) | 172 (137 to 212) | 202 (175 to 232) | -30 (-65 to 10) |
| Other males |  |  |  |  |  |
|  | Chlamydia | Non-recipients (indirect effect) | 7,815 (6,364 to 9,664) | 7,765 (7,593 to 7,940) | 50 (-1,401 to 1,899) |
|  | Syphilis | Non-recipients (indirect effect) | 616 (418 to 922) | 585 (539 to 634) | 31 (-167 to 337) |
|  | Gonorrhea | Non-recipients (indirect effect) | 2,329 (1,775 to 3,062) | 2,345 (2,251 to 2,442) | -16 (-570 to 717) |
| Females |  |  |  |  |  |
|  | Chlamydia | Non-recipients (indirect effect) | 16,419 (13,583 to 20,076) | 16,990 (16,735 to 17,247) | -571 (-3,407 to 3,086) |
|  | Syphilis | Non-recipients (indirect effect) | 281 (173 to 474) | 294 (261 to 330) | -13 (-121 to 180) |
|  | Gonorrhea | Non-recipients (indirect effect) | 1,534 (1,175 to 2,012) | 1,896 (1,812 to 1,983) | -362 (-721 to 116) |

CI: Confidence interval, estimated via bootstrap resampling with 10,000 iterations. We derive estimates of expected cases, absent overall or indirect effects, via time-series analyses projecting a continuation of pre-implementation slopes for each population (**Figure 2**). We derive estimates of expected cases, absent direct effects, via the inverse of the adjusted odds ratio of recent (<90d) doxyPEP receipt among individuals testing positive or negative for each bSTI (**Table 3**). We multiply incidence of each bSTI within 90d after doxyPEP dispenses, among doxyPEP recipients,via the inverse of the adjusted odds ratio to obtain expected cases, absent direct effects.
<sup>1</sup>We define at-risk MSM and TGW as those living with HIV or those with any history of receipt of HIV pre- or post-exposure prophylaxis from July 1, 2021 onward. Other males were HIV negative without history of pre- or post-exposure prophylaxis receipt after July 1, 2021.

**Table S9:** Indirect and total effects of doxyPEP among at-risk MSM and TGW, applying a 180-day exposure history definition.

| **Outcome** | **Population** | **Incidence rate, cases per 1,000 person-years (95% CI)** |  | **Estimate (95% CI)** |  |
|---|---|---|---|---|---|
|  |  | Observed | Expected<sup>1</sup> | Effect, %<sup>2</sup> | Incidence averted, cases per 1,000 person-years |
| Chlamydia |  |  |  |  |  |
|  | Indirect effect | 81.3 (78.7 to 84.0) | 135.2 (118.1 to 155.4) | 40.1 (31.3 to 47.9) | 53.9 (36.8 to 74.0) |
|  | Total effect | 46.1 (35.5 to 58.8) | 221.8 (179.8 to 270.4) | 75.2 (67.3 to 81.3) | 175.8 (133.8 to 224.4) |
| Syphilis |  |  |  |  |  |
|  | Indirect effect | 21.7 (20.3 to 23.1) | 29.9 (24.4 to 36.9) | 28.0 (11.6 to 41.4) | 8.3 (2.8 to 15.2) |
|  | Total effect | 8.6 (4.5 to 15.1) | 35.1 (23.4 to 50.2) | 70.5 (45.7 to 83.9) | 26.4 (14.8 to 41.6) |
| Gonorrhea |  |  |  |  |  |
|  | Indirect effect | 118.6 (115.4 to 121.8) | 118.4 (104.2 to 134.6) | 0.3 (-13.8 to 12.3) | -0.1 (-14.4 to 16.0) |
|  | Total effect | 226.7 (202.4 to 253.2) | 195.9 (160.1 to 238.1) | -19.4 (-42.0 to -0.5) | -30.8 (-66.6 to 11.4) |

CI: Confidence interval, estimated via bootstrap resampling with 10,000 iterations. We define at-risk MSM and TGW as those living with HIV or those with any history of receipt of HIV pre- or post-exposure prophylaxis from July 1, 2021 onward.
<sup>1</sup>We estimate expected incidence rates via interrupted time-series analyses excluding post-intervention changes in slopes, for incidence rates of each infection (**Figure 2**). Estimates among individuals who did not receive doxyPEP (indirect effects) exclude cases occurring within 90 days after doxyPEP dispenses.
<sup>2</sup>Estimated effects are reported as observed cases divided by expected cases.

**Table S10:**
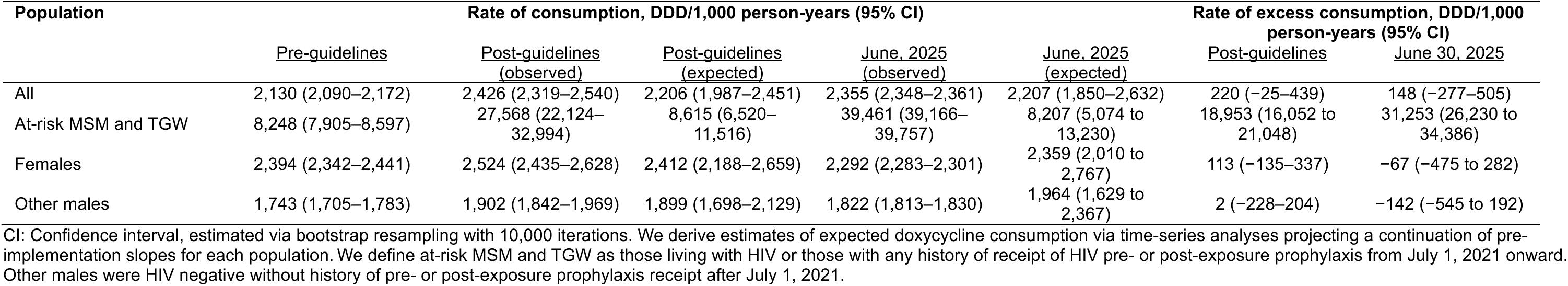
Change in doxycycline consumption after doxyPEP implementation, by population stratum.

| Population | Rate of consumption, DDD/1,000 person-years (95% CI) |  |  |  |  | Rate of excess consumption, DDD/1,000 person-years (95% CI) |  |
|---|---|---|---|---|---|---|---|
|  | Pre-guidelines | Post-guidelines (observed) | Post-guidelines (expected) | June, 2025 (observed) | June, 2025 (expected) | Post-guidelines | June 30, 2025 |
| All | 2,130 (2,090–2,172) | 2,426 (2,319–2,540) | 2,206 (1,987–2,451) | 2,355 (2,348–2,361) | 2,207 (1,850–2,632) | 220 (−25–439) | 148 (−277–505) |
| At-risk MSM and TGW | 8,248 (7,905–8,597) | 27,568 (22,124–32,994) | 8,615 (6,520–11,516) | 39,461 (39,166–39,757) | 8,207 (5,074 to 13,230) | 18,953 (16,052 to 21,048) | 31,253 (26,230 to 34,386) |
| Females | 2,394 (2,342–2,441) | 2,524 (2,435–2,628) | 2,412 (2,188–2,659) | 2,292 (2,283–2,301) | 2,359 (2,010 to 2,767) | 113 (−135–337) | −67 (−475 to 282) |
| Other males | 1,743 (1,705–1,783) | 1,902 (1,842–1,969) | 1,899 (1,698–2,129) | 1,822 (1,813–1,830) | 1,964 (1,629 to 2,367) | 2 (−228–204) | −142 (−545 to 192) |

CI: Confidence interval, estimated via bootstrap resampling with 10,000 iterations. We derive estimates of expected doxycycline consumption via time-series analyses projecting a continuation of pre-implementation slopes for each population. We define at-risk MSM and TGW as those living with HIV or those with any history of receipt of HIV pre- or post-exposure prophylaxis from July 1, 2021 onward. Other males were HIV negative without history of pre- or post-exposure prophylaxis receipt after July 1, 2021.

**Table S11:** Absolute rate of doxycycline consumption, by population stratum.

| Population | Rate, total DDD/year (95% CI) |  |  |
|---|---|---|---|
|  | January 1, 2022 to April 27, 2023 | April 28, 2023 to June 30, 2025 | Rate as of June 30, 2025 |
| All | 5,924,572 (5,869,097–5,980,044) | 6,621,594 (6,571,412–6,671,343) | 6,501,426 (6,344,415–6,660,123) |
| At-risk MSM and TGW | 172,708 (163,896–181,819) | 586,991 (573,533–600,675) | 833,793 (791,643–876,150) |
| Females | 3,455,587 (3,413,896–3,496,824) | 3,584,420 (3,549,905–3,619,953) | 3,295,931 (3,183,106–3,411,502) |
| Other males | 2,296,277 (2,260,951–2,332,656) | 2,450,183 (2,418,279–2,481,925) | 2,371,702 (2,273,706–2,471,690) |

We derive estimates of expected doxycycline consumption via time-series analyses projecting a continuation of pre-implementation slopes for each population. We define at-risk MSM and TGW as those living with HIV or those with any history of receipt of HIV pre- or post-exposure prophylaxis from July 1, 2021 onward. Other males were HIV negative without history of pre- or post-exposure prophylaxis receipt after July 1, 2021.

**Table S12:**
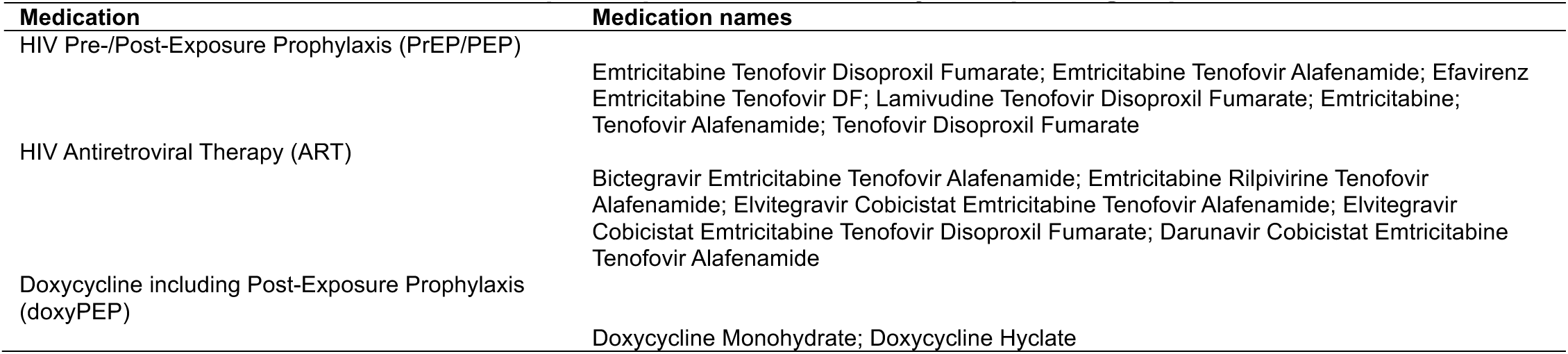
Classification of HIV-related prescription medications by therapeutic group.

| Medication | Medication names |
|---|---|
| HIV Pre-/Post-Exposure Prophylaxis (PrEP/PEP) |  |
|  | Emtricitabine Tenofovir Disoproxil Fumarate; Emtricitabine Tenofovir Alafenamide; Efavirenz Emtricitabine Tenofovir DF; Lamivudine Tenofovir Disoproxil Fumarate; Emtricitabine; Tenofovir Alafenamide; Tenofovir Disoproxil Fumarate |
| HIV Antiretroviral Therapy (ART) |  |
|  | Bictegravir Emtricitabine Tenofovir Alafenamide; Emtricitabine Rilpivirine Tenofovir Alafenamide; Elvitegravir Cobicistat Emtricitabine Tenofovir Alafenamide; Elvitegravir Cobicistat Emtricitabine Tenofovir Disoproxil Fumarate; Darunavir Cobicistat Emtricitabine Tenofovir Alafenamide |
| Doxycycline including Post-Exposure Prophylaxis (doxyPEP) |  |
|  | Doxycycline Monohydrate; Doxycycline Hyclate |

**Table S13:**
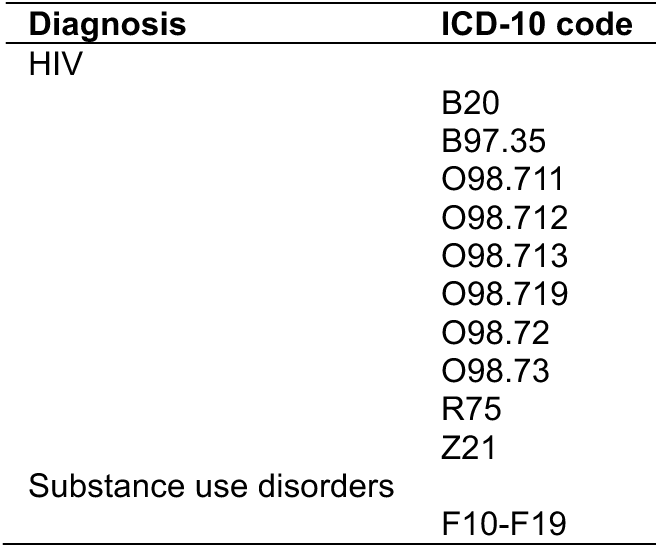
ICD-10 codes of STI-related diagnoses.

**Table S14:** Indirect effects among females and other males in analyses accounting for change in intercept at doxyPEP implementation.

| **Population** | **Outcome** | **Incidence rate, cases per 1,000 person-years (95% CI)** |  | **Estimate (95% CI)** |  |
|---|---|---|---|---|---|
|  |  | Observed | Expected | Effect, % | Incidence averted, cases per 1,000 person-years |
| Other males<sup>1</sup> |  |  |  |  |  |
|  | Chlamydia | 2.8 (2.7 to 2.8) | 3.1 (2.7 to 3.6) | 9.9 (–4.5 to 22.4) | 0.3 (–0.1 to 0.8) |
|  | Syphilis | 0.2 (0.2 to 0.2) | 0.2 (0.2 to 0.3) | 14.6 (–12.6 to 35.8) | 0.0 (–0.0 to 0.1) |
|  | Gonorrhea | 0.8 (0.8 to 0.9) | 0.8 (0.7 to 1.0) | –3.9 (–25.4 to 15.3) | 0.0 (–0.2 to 0.2) |
| Females |  |  |  |  |  |
|  | Chlamydia | 5.5 (5.4 to 5.6) | 5.9 (5.1 to 6.7) | 6.1 (–7.6 to 18.3) | 0.4 (–0.4 to 1.2) |
|  | Syphilis | 0.1 (0.1 to 0.1) | 0.1 (0.1 to 0.1) | 6.9 (–32.0 to 36.2) | 0.0 (0.0 to 0.1) |
|  | Gonorrhea | 0.6 (0.6 to 0.6) | 0.5 (0.4 to 0.6) | –32.1 (–58.7 to –8.8) | –0.1 (–0.2 to 0.0) |

CI: Confidence interval, estimated via bootstrap resampling with 10,000 iterations. We derive estimates of expected cases, absent overall or indirect effects, via time-series analyses projecting a continuation of pre-implementation slopes for each population (**Figure 2**) additionally accounting for post-implementation changes in intercepts as a component of intervention effects.
<sup>1</sup>Males included in these analyses exclude those living with HIV or those with any history of receipt of HIV pre- or post-exposure prophylaxis from July 1, 2021 onward.

**Table S15:**
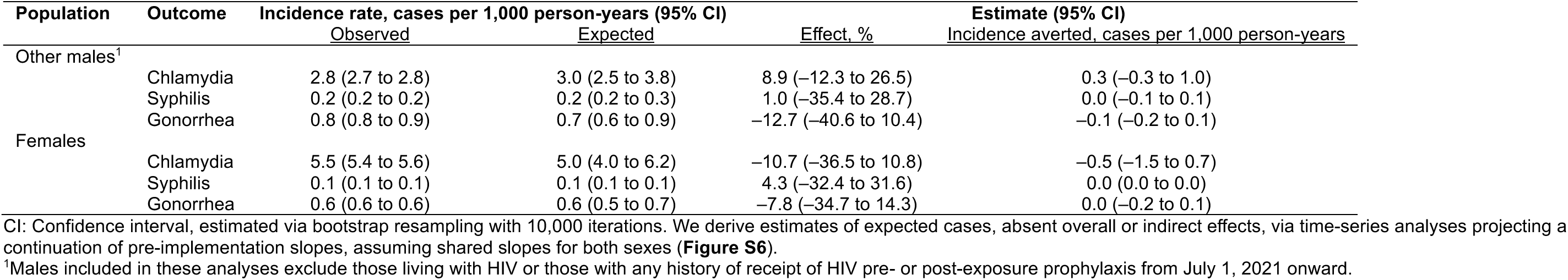
Indirect effects among males not classified as at-risk and females in analyses assuming shared slopes.

| Population | Outcome | Incidence rate, cases per 1,000 person-years (95% CI) |  | Estimate (95% CI) |  |
|---|---|---|---|---|---|
|  |  | Observed | Expected | Effect, % | Incidence averted, cases per 1,000 person-years |
| Other males<sup>1</sup> |  |  |  |  |  |
|  | Chlamydia | 2.8 (2.7 to 2.8) | 3.0 (2.5 to 3.8) | 8.9 (–12.3 to 26.5) | 0.3 (–0.3 to 1.0) |
|  | Syphilis | 0.2 (0.2 to 0.2) | 0.2 (0.2 to 0.3) | 1.0 (–35.4 to 28.7) | 0.0 (–0.1 to 0.1) |
|  | Gonorrhea | 0.8 (0.8 to 0.9) | 0.7 (0.6 to 0.9) | –12.7 (–40.6 to 10.4) | –0.1 (–0.2 to 0.1) |
| Females |  |  |  |  |  |
|  | Chlamydia | 5.5 (5.4 to 5.6) | 5.0 (4.0 to 6.2) | –10.7 (–36.5 to 10.8) | –0.5 (–1.5 to 0.7) |
|  | Syphilis | 0.1 (0.1 to 0.1) | 0.1 (0.1 to 0.1) | 4.3 (–32.4 to 31.6) | 0.0 (0.0 to 0.0) |
|  | Gonorrhea | 0.6 (0.6 to 0.6) | 0.6 (0.5 to 0.7) | –7.8 (–34.7 to 14.3) | 0.0 (–0.2 to 0.1) |

CI: Confidence interval, estimated via bootstrap resampling with 10,000 iterations. We derive estimates of expected cases, absent overall or indirect effects, via time-series analyses projecting a continuation of pre-implementation slopes, assuming shared slopes for both sexes (**Figure S6**).
<sup>1</sup>Males included in these analyses exclude those living with HIV or those with any history of receipt of HIV pre- or post-exposure prophylaxis from July 1, 2021 onward.

**Figure S1:**
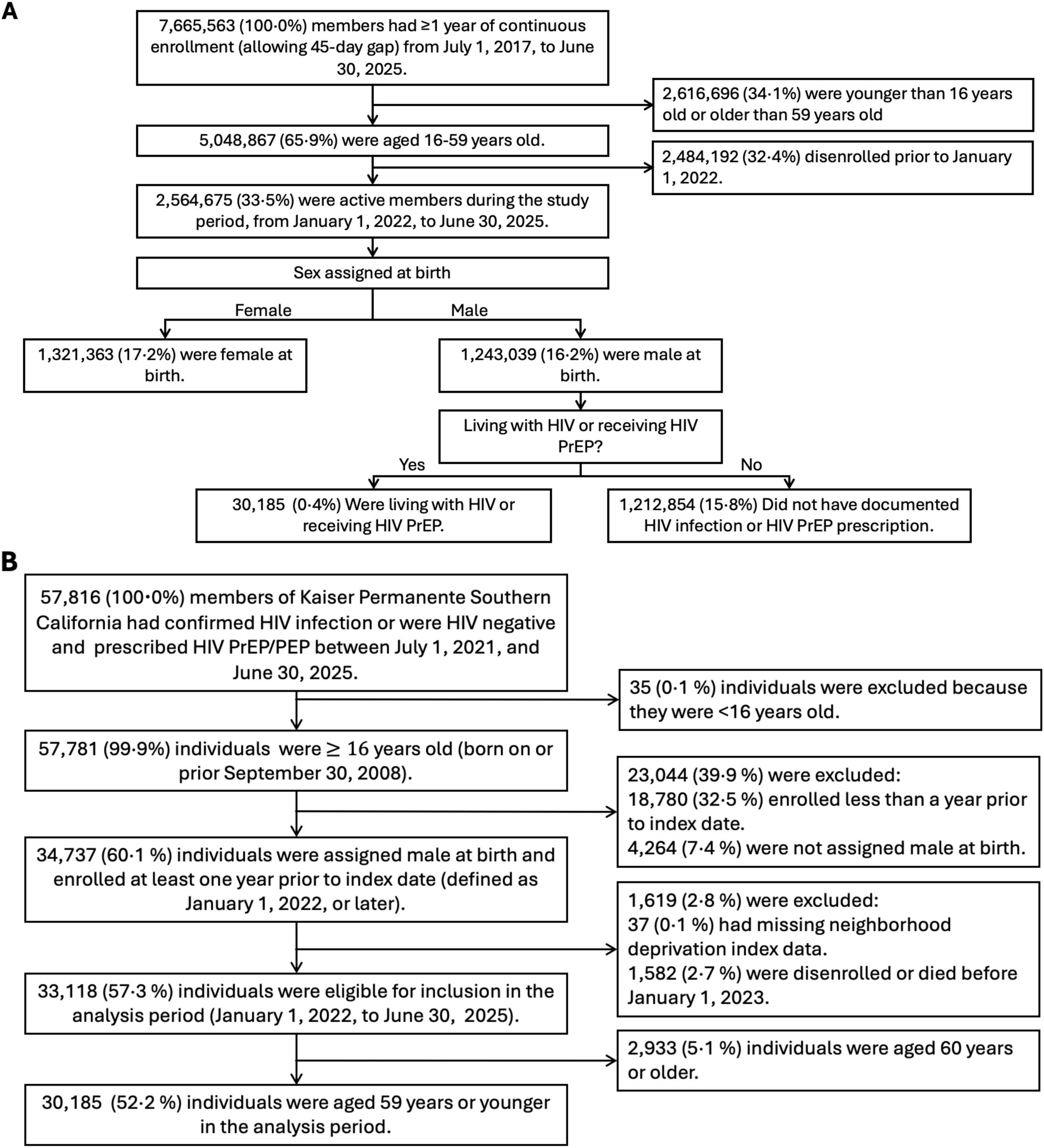
Study flowchart. We detail the numbers of individuals meeting eligibility criteria and included in the study as (**a**) females and males without confirmed HIV infection or an HIV PrEP/PEP prescription and (**b**) “at-risk” males living with HIV or receiving HIV PrEP/PEP.

**Figure S2:**
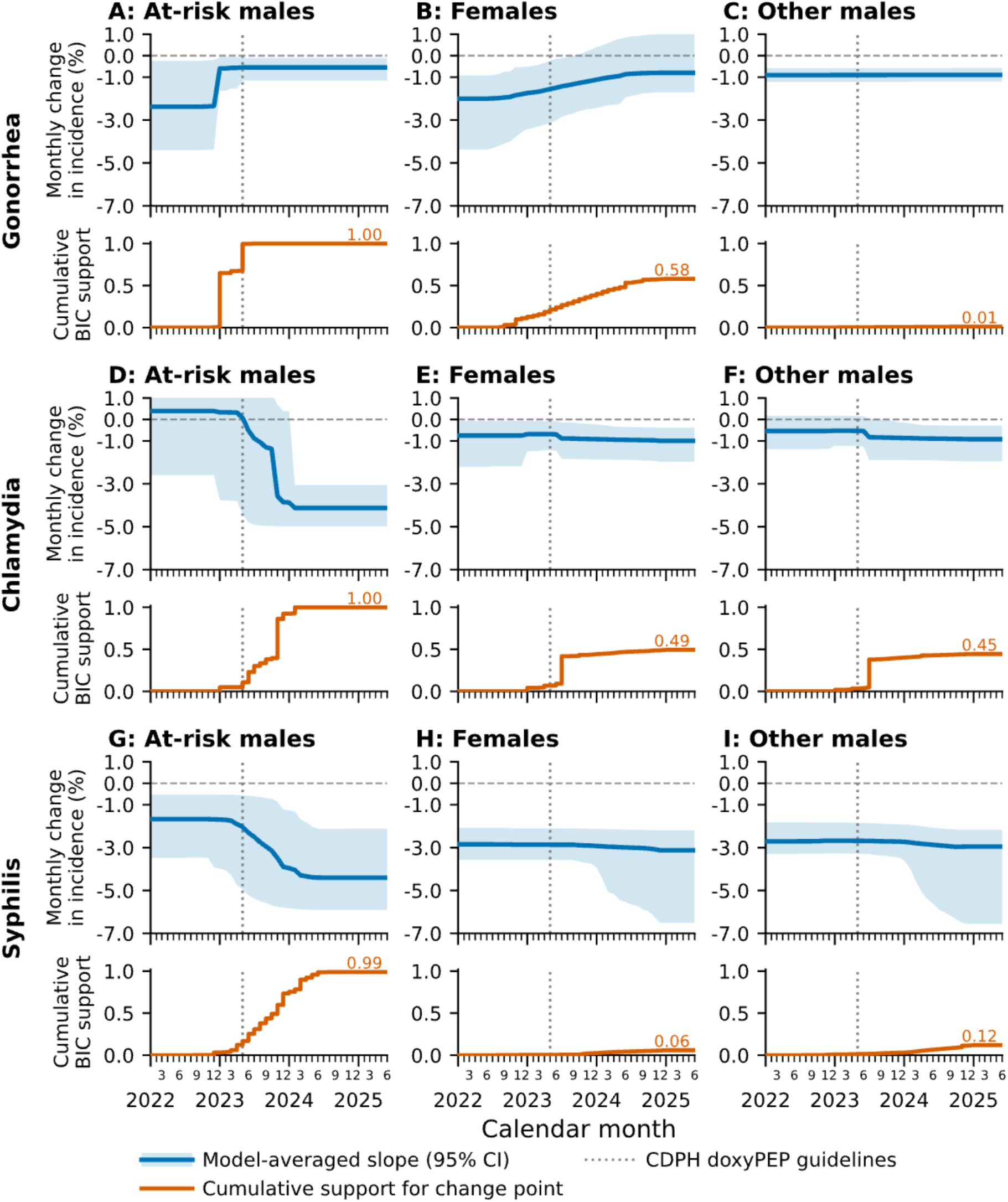
Model-averaged monthly incidence slopes and cumulative support for changepoints identified by breakpoint analysis. We illustrate the BIC-weighted average monthly incidence slope across models with different candidate changepoints, including a no-change model (top), together with the cumulative model support that a change point had occurred by month t (bottom). Results are stratified by organism (**a-c**) gonorrhea, (**d-f**) chlamydia, and (**g-i**) syphilis, and by population group: (**a, d, g**) at-risk males (living with HIV or with a history of receiving HIV PrEP/PEP), (**b, e, h**) females, and (**c, f, i**) other males. All models included calendar time as a continuous variable and were adjusted for age group and seasonal variation. Shaded areas represent 95% confidence intervals.

**Figure S3:**
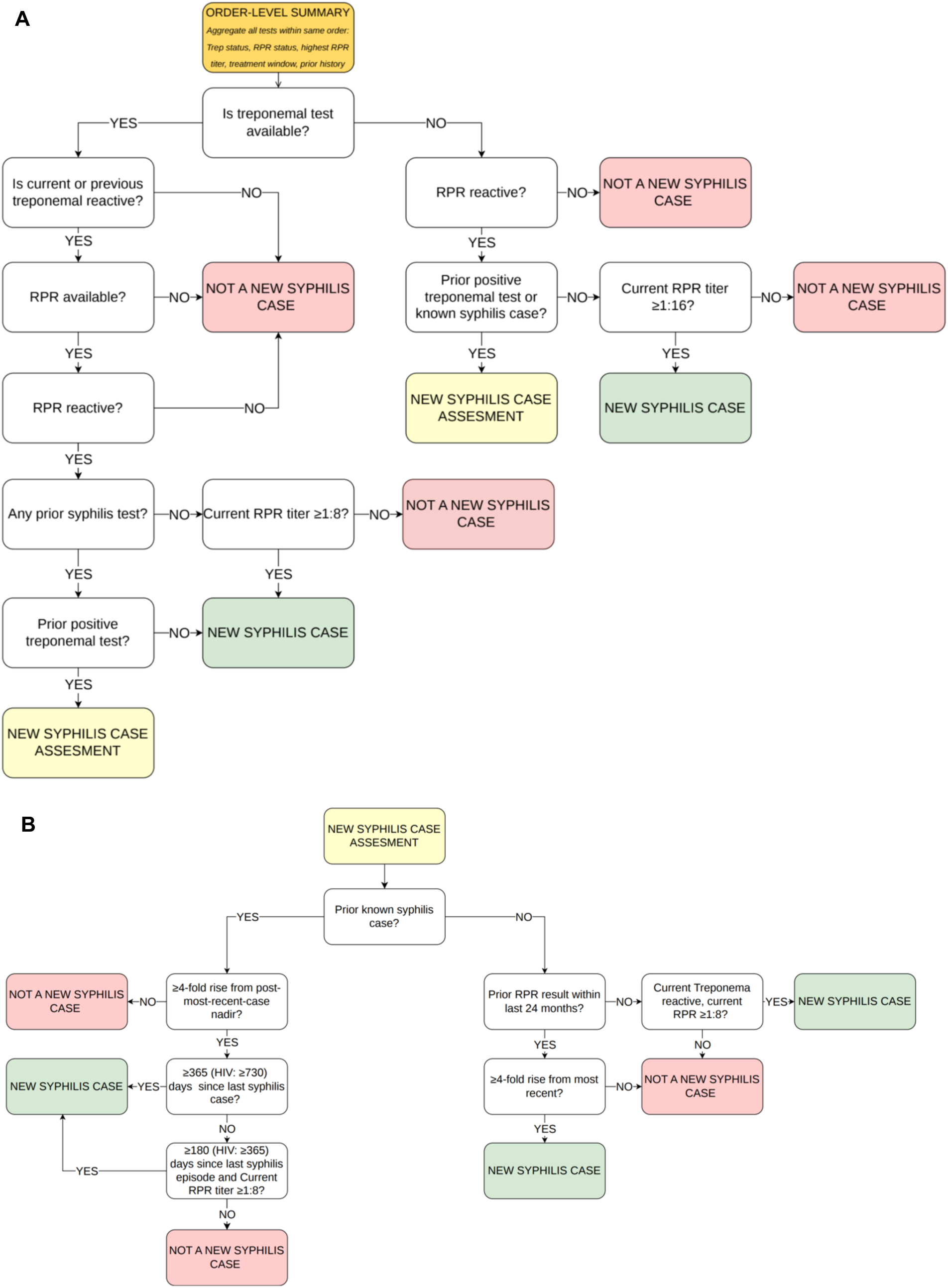
Flowchart of the algorithm for identifying new syphilis infections from longitudinal laboratory data. We detail the algorithm used to classify incident syphilis infections from laboratory testing data, including (**a**) the initial phase to identify cases requiring further assessment for a new syphilis infection and (**b**) the criteria applied to those cases. If the most recent prior RPR result was nonreactive, we defined a significant rise as a current RPR titer of ≥1:4. We defined the nadir in titers after the most recent infection as the lowest RPR titer recorded following the most recent syphilis infection.

**Figure S4:**
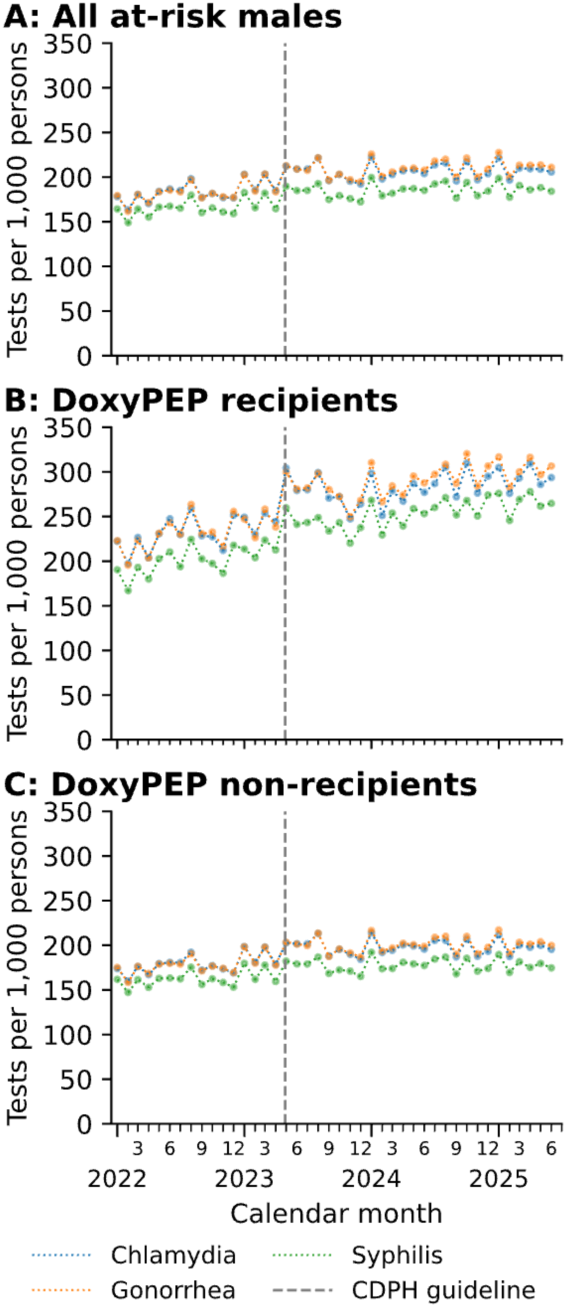
Observed monthly testing rates for gonorrhea, chlamydia, and syphilis among at-risk males. We illustrate monthly laboratory testing rates per 1,000 persons of chlamydia, gonorrhea, and syphilis among at-risk males living with HIV or receiving HIV PrEP/PEP stratified by (**a**) all individuals, (**b**) individuals who ever received doxyPEP, and (**c**) individuals who never received doxyPEP throughout the study period.

**Figure S5:**
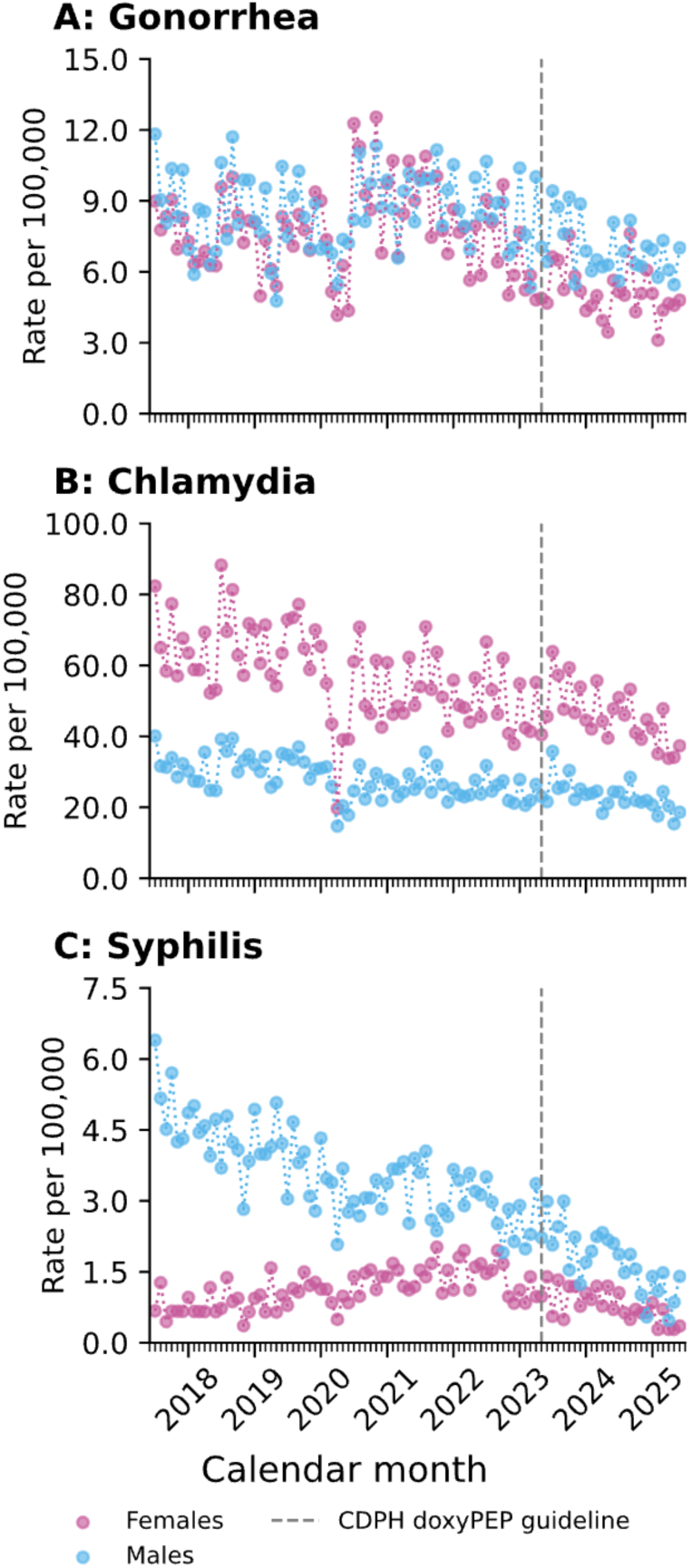
Extended incidence time series for gonorrhea, chlamydia, and syphilis diagnoses among females and males outside the at-risk population. We illustrate monthly incidence rates per 100,000 persons for (a) gonorrhea, (b) chlamydia, and (c) syphilis from July 2017 through June 2025 among females and males not belonging to the at-risk population (HIV-negative without history of PrEP/PEP receipt).

**Figure S6:**
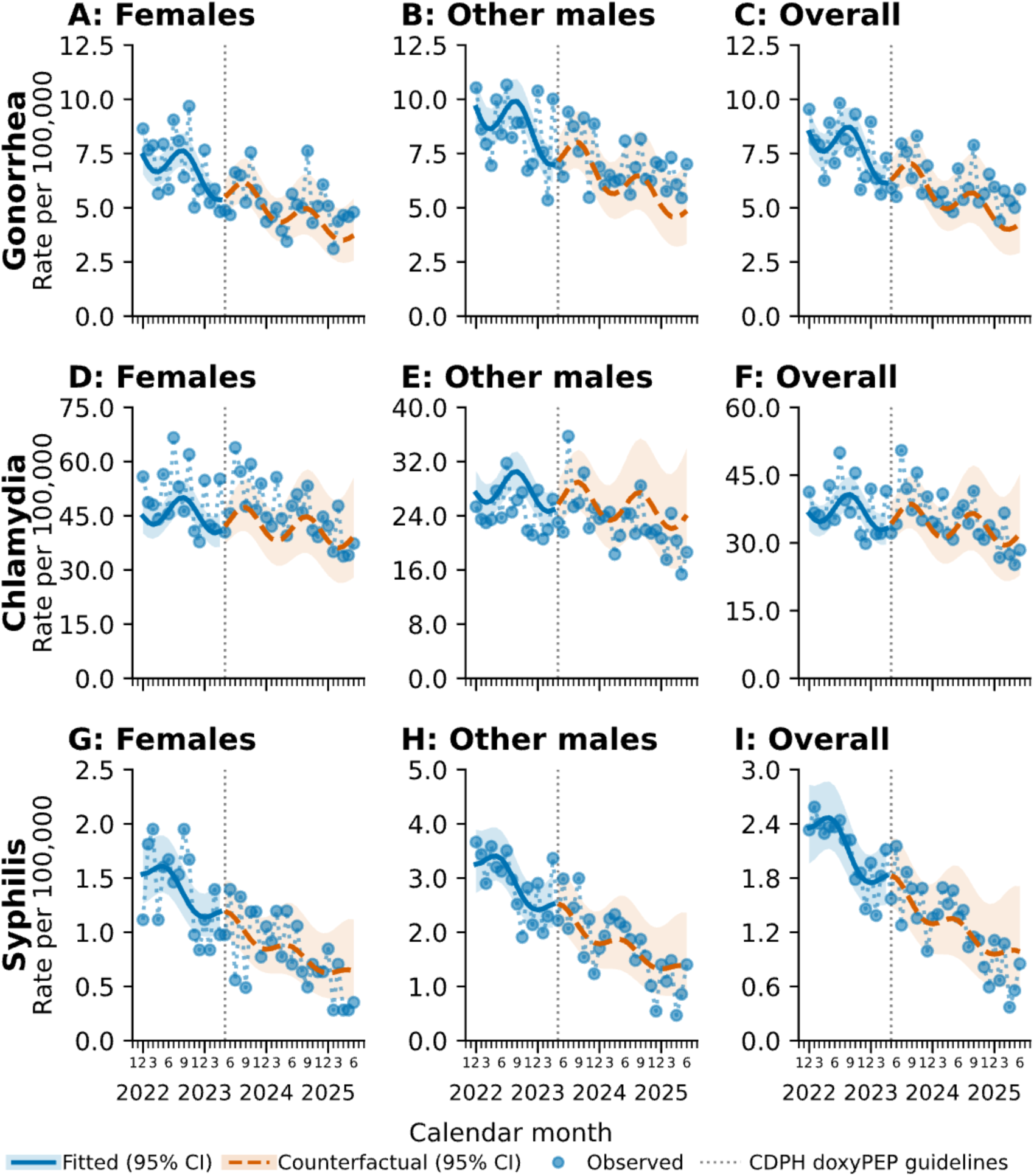
Observed and counterfactual trends in gonorrhea, chlamydia, and syphilis incidence following doxyPEP guideline implementation in analyses assuming shared slopes. Analyses resemble those illustrated in Figure 2 but define shared slope parameters for females and other males (HIV-negative without history of PrEP/PEP receipt), assuming similar trends in bSTI incidence within heterosexual networks.

## Notes

### Competing Interest Statement

The authors have declared no competing interest.

### Summary of Updates

Updated analyses for quantification of direct, indirect, total, and overall effects. Updated introduction and discussion addressing context of "core group" transmission dynamics.

